# Rationale and design of the Health Professional Students at the University of Illinois Chicago (HOLISTIC) Cohort Study

**DOI:** 10.1101/2021.11.12.21266272

**Authors:** Sunil R. Dommaraju, Stephanie Gordon Rivera, Ethan G. Rocha, Scott Bicknell, Daniel Loizzo, Ayesha Mohammad, Priya Rajan, Alexandria Seballos, Avisek Datta, Rashid Ahmed, Jerry A. Krishnan, Mary Keehn

## Abstract

**Objectives:** The objectives of the HOLISTIC Cohort Study are to establish a 3-year prospective cohort study that characterizes the health of students within and across health professionals’ education programs during the coronavirus disease 2019 (COVID-19) pandemic, implement an interprofessional student research team, and inform initiatives to improve student health. This report describes the rationale and design of the HOLISTIC Cohort Study, including recruitment strategy, survey development, data management, and descriptive statistics of the first wave of study participants.

**Methods:** An interprofessional student research team was formed to continuously inform study design. The first wave of recruitment was conducted from April 14, 2021 to May 5, 2021 across seven health science colleges (applied health, dentistry, medicine, nursing, pharmacy, public health, social work) at the University of Illinois Chicago in Chicago, IL. Eligible students were sent an invitation via email to complete an online survey after providing electronic informed consent. The online survey was based on the U.S. Centers for Disease Control and Prevention’s Behavioral Risk Factor Surveillance System 2019 survey and the 2014 World Health Organization Report of the Strategic Advisory Group of Experts Working Group Questionnaire. Two additional recruitment waves are planned in the Spring 2022 and Spring 2023; follow-up of participants previously enrolled will occur during these second and third recruitment waves.

**Results:** Of 5,118 students invited to participate in the first wave, 553 (10.8%) completed the survey and includes participants from all seven health science colleges. The average age of participants is 27.3 years, 435 (78.8%) identify as female, and 137 (24.8%) identify as an underrepresented minority. Overall, 465 (84.6%) participants reported being currently employed for wages. Just over half (51%) reported no days with poor physical health within a month but only 11.2% reported no days with poor mental health within a month. Nearly one in ten (9.4%) reported having ever had a positive test for COVID-19.

**Conclusion:** The HOLISTIC Cohort Study of health professional students across seven health science colleges has completed the first of three waves of enrollment during the COVID-19 pandemic. Based on the first wave of study participants, increased attention to supporting the mental and physical health of health professional students is needed.

## INTRODUCTION

Prospective cohort studies examining health professionals have been crucial to our understanding of health determinants and have directly shaped healthcare interventions. For example, the British Doctors Study investigated the effects of smoking tobacco on mortality, myocardial infarction, lung cancer, and lung disease, ^1^ the Nurses’ Health Studies established recommendations regarding women’s health, ^2^ the Physicians’ Health Study established guidelines to prevent cardiovascular disease and cancer, ^3^ and the Health Professionals Follow-Up Study has made numerous conclusions about the health of male dentists, pharmacists, optometrists, osteopath physicians, podiatrists, and veterinarians.^4^

Cross-sectional studies of *students* in the health professions have similarly examined various health-related topics, including health literacy,^5^ use of e-cigarettes,^6^ empathy,^7^ and mental health.^8^ In 2020, particular attention has been paid to how the coronavirus disease 2019 (COVID-19) pandemic has affected the didactic and experiential learning environment of health professional students. A National Academies of Sciences, Engineering, and Medicine workshop report in 2020 highlighted the grand challenges facing health professions education (HPE) during the COVID-19 pandemic.^9^ For example, clinical training of dental students temporarily relied on case presentations, rather than experiential learning alongside practicing clinicians due to infection control precautions.^10^ Another report found that 70% of medical students reported worsening mental health during the initial months of the pandemic.^11^

Prospective cohort studies of health professional students initiated during the COVID-19 pandemic could provide much needed information about changes in the physical, mental, and social health of students during the pandemic and their occupational trajectory as they enter the health professional workforce. These insights could inform the design and evaluation of interventions to promote student health and HPE during and beyond the COVID-19 pandemic.

The Health Professional Students at the University of Illinois Chicago (HOLISTIC) Cohort Study enrolled and will prospectively track student participants across seven health science colleges (applied health sciences, dentistry, medicine, nursing, pharmacy, public health, and social work) at the University of Illinois Chicago (UIC), an urban, 4-year, public university in Chicago, Illinois, USA. The primary objective of the HOLISTIC Cohort Study is to establish a prospective cohort study that characterizes the health and health behaviors of students within and across health professionals’ education programs during the COVID-19 pandemic. Additional objectives are to develop the interprofessional collaboration skills of students through an interprofessional mentored student research team and to inform initiatives to support and improve the health and health behaviors of health professions students. The HOLISTIC Cohort Study may also provide insight into changes in health and health behaviors as graduates enter the workforce and some early indicators of occupational trajectories across the health sciences.

The purpose of this report is to describe the rationale and design of the HOLISTIC Cohort Study, including recruitment strategy, survey development, data management, and basic descriptive statistics of the first wave of participants. Lessons learned during initial stages of study implementation are also discussed.

## METHODS

### Overview

The HOLISTIC Cohort Study was designed to enroll and prospectively track the health and health behaviors of students across the health professions for a minimum of three years. The UIC Institutional Review Board approved the study (protocol #2021-0114). The Interprofessional Practice and Education (IPE) program within the UIC Office of Vice Chancellor for Health Affairs (OVCHA) provided overall project oversight.

The design and implementation of the HOLISTIC Cohort Study was supported by faculty mentors in three health science colleges (MK, Assistant Vice Chancellor for IPE and Associate Dean for Clinical Affairs in the College of Applied Health sciences; JK, Associate Vice Chancellor for Population Health Sciences, Professor in the College of Medicine; and RA, Associate Dean for Academic Affairs, Professor in the School of Public Health). The initial focus was on the development of an interprofessional student research team to design and implement the HOLISTIC Cohort Study. The faculty mentors worked closely with deans and faculty of each UIC health science college to receive continuous feedback on and develop improvements for the design of the study.

### Recruitment of student researchers

The formation of the HOLISTIC Cohort Study’s student research team began with the recruitment of two student leaders from the College of Medicine and College of Applied Health Sciences who formed the administrative workgroup of the student research team. These two students then began recruiting other health professional students across all seven health science colleges at UIC who were interested in conducting this type of research through an interest form that was emailed to all health professional students. The goal was to have two to three student liaisons per health science college.

The student research team was comprised of an interprofessional group representing the following health science colleges: College of Pharmacy, School of Public Health, College of Nursing, College of Applied Health Science, College of Dentistry, and College of Medicine. Forming an interprofessional team was crucial to the success of the project since each team member brought a unique perspective from the lens of their health profession. These perspectives helped inform development of all aspects of the study methodology. The student research team developed a collaborative practice through innovation and learning during the process.

The team was organized into five workgroups with defined responsibilities (administrative, recruitment and retention, external communication, survey, and regulatory), as shown in **Table 1**, based on student preferences and to ensure that each workgroup included students from at least two health science colleges.

**Table 1:** Organization of the HOLISTIC cohort study research team.

| **Workgroup** | **Role** |
| --- | --- |
| Administrative | Serve as liaisons to coordinate tasks and timelines among workgroups, facilitate communication with UIC stakeholders, and act as project managers |
| Recruitment & Retention | Create recruitment materials such as email messages, flyers, study overview document, and compile listservs for each eligible health science program |
| External Communication | Curate the HOLISTIC Cohort Study website and collaborate with deans of each health science college to communicate study needs |
| Regulatory | Prepare and manage IRB materials, develop necessary amendments, and create future submissions |
| Survey | Organize the survey questionnaire and build the online REDCap infrastructure |

### Study participant eligibility criteria

To be eligible to participate, students were required to be 1) age 18 years or older; 2) enrolled full- or part-time in one or more of UIC’s seven health science colleges; and 3) enrolled in a program that prepares its graduates to enter a healthcare profession. The list of eligible health science degree programs is included in **Appendix 3**.

### Recruitment of study participants

Enrollment into the HOLISTIC Cohort Study is designed to occur during a series of three recruitment waves (Spring 2021 [April 14, 2021 to May 5, 2021; completed], Spring 2022, Spring 2023). To ensure representation of students across all the health science colleges, the team sought to enroll a convenience sample of students in each health science college that was proportional to the number of part- and full-time students in each college; the total goal recruitment number was set at 2,000 participants based on available resources such as the enrollment window and funds to support reimbursement of study participants (**Table 2**).

**Table 2:** Target enrollment, by UIC health science college.

| University of Illinois Chicago,<br>Health Science College | Students enrolled part- or full-time,<br>Fall 2020, number | Eligible students, number | Target enrollment, number |
| --- | --- | --- | --- |
| College of Applied Health Sciences | 2,111 | 462 | 181 |
| College of Medicine | 1,487 | 1,264 | 494 |
| College of Dentistry | 473 | 378 | 148 |
| College of Nursing | 1,504 | 1,398 | 546 |
| College of Pharmacy | 863 | 742 | 290 |
| School of Public Health | 821 | 396 | 155 |
| Jane Addams College of Social Work | 499 | 478 | 186 |
| **Total** | **7,758** | **5118** | **2,000** |

**Footer:** The University of Illinois Chicago (UIC) includes seven health sciences colleges. In the Fall 2020, there were a total of 7,758 part- and full-time students enrolled across the health sciences colleges, of which 5,118 (66%) were eligible to participate in the HOLISTIC study (see Methods; Source Fall 2020 UIC OIR Census File). The target enrollment in the HOLISTIC study is 2,000 health sciences students, representing about 39% of eligible students in each college.

Recruitment procedures were designed to minimize in-person interactions between the target population of students and the student research team to reduce the risk of COVID-19 exposure, the risk of coercion or undue influence that may occur among peers, and the possibility for social desirability bias when providing responses to survey items. Thus, the study was administered using Research Electronic Data Capture (REDCap), a secure, web-based software platform designed to support data capture for research studies.^12^ For this study, the REDCap platform included all recruitment materials, prescreening questions, electronic informed consent (e-consent), and the HOLISTIC Cohort Study survey.

The following steps were employed to recruit subjects:

1. *Recruitment emails:* Students in eligible programs received an email sent by their education program’s listserv administrator detailing the study, eligibility requirements, and providing a link inviting them to learn more about the study (study overview). The survey was active for 14 days from when the subject received the initial recruitment email. There were two reminder emails, one that was sent 5 days after the initial recruitment email and a second, final, reminder email that was sent two days before the survey was to close. Recruitment was staged one college at a time over a 3-week period to allow for the student research team to troubleshoot survey issues early in the recruitment process.
2. *Study overview:* The overview includes the study’s purpose, procedures, potential risks and benefits, contact information for questions, and a link to guide participants to the screening questions.
3. *Screening questions:* While the initial recruitment email was only sent to eligible students, two screening questions were required to confirm that participants are 18 years or older and are enrolled in at least one of the eligible degree programs based on self-report. Eligible participants were directed to a e-consent document on REDCap.
4. *E-consent:* Subjects were invited to read IRB-approved text in English and provide e-consent to participate in the study.
5. *Survey completion:* Participants who offered e-consent were directed to the survey. After completing the survey, participants received an email with a $5 e-gift card as compensation for the time spent completing the survey.

### Retention

A multi-pronged strategy will be used to promote retention of study participants, including: 1) collecting full name, email address, and phone number of study participants for future follow-up 2) presenting at student events, and 3) sharing information and preliminary, de-identified results on a study website.^13^ Further input from the interprofessional student research team will be crucial to identify retention strategies, including college-specific strategies. Input from participants in the health science colleges will be sought as well. The HOLISTIC Cohort Study is designed to follow study participants for up to three years after enrollment.

### Survey development

The HOLISTIC Cohort Study collected participant contact information, participant demographics, and responses to questions about health and health related behaviors including vaccination (**Table 3**). Completion of the survey was anticipated to take about 15 to 20 minutes based on pilot testing by the student research team; the survey was administered via REDCap.^12^

**Table 3:** HOLISTIC Cohort Study survey domains.

| **CATEGORY** | **DOMAINS** |
| --- | --- |
| **General Information** | Students' identifiers, UIC health science college enrollment, degree program enrollment |
| **Demographics** | Sex at birth, date of birth, race/ethnicity, marital status, education level, residence, zip code, housing arrangement, type of telephone number, cellphones, military service, employment status, number of children leaving in the household, annual income, weight, height, pregnant, deaf, blind, difficulty concentrating, difficulty walking, difficulty dressing, difficulty doing errands |
| **Health status and health-related behaviors** | Health status, healthy days, healthcare access, hypertension awareness, cholesterol awareness, chronic health conditions, tobacco use, alcohol consumption, exercise, fruits & vegetable consumption, immunizations, prediabetes, diabetes, vaccinations, breast & cervical cancer screening, colorectal cancer screening, adverse childhood experiences, marijuana use, sexual orientation & gender identity |

The survey was based on two validated questionnaires: 1) the U.S. Centers for Disease Control and Prevention’s (CDC) Behavioral Risk Factor Surveillance System (BRFSS) 2019 survey,^14^ and 2) the 2014 World Health Organization Report of the Strategic Advisory Group of Experts (WHO SAGE) Working Group Questionnaire.^15,16^ Using a deliberative process, the student research team and faculty mentors identified 12 of 14 core sections of the BRFSS for inclusion in the survey, removing “Arthritis” and “HIV/AIDS” core sections due to concerns about relevance to target population and the need for additional documentation and consent from participants, respectively. The student research team and faculty mentors also elected to include 10 of 31 optional BRFSS modules that reflect national leading health indicators as established by programs such as Healthy People 2020 or the Chicago Health Survey (**Table 3**).^17,18^ The 2014 WHO SAGE Working Group questionnaire was used to examine vaccine hesitancy among health professional students.^15,16^ Vaccine hesitancy is a barrier to controlling the spread of COVID-19, and the WHO SAGE Working Group questionnaire has been used to estimate the rate of vaccine hesitancy across the globe.

The student research team made minor modifications to the instructions, wording of the questions, and response options of the BRFSS and WHO SAGE Working Group questionnaire for clarity of administration via an online portal, rather than telephone administration as originally intended by BRFSS. The team did not modify the original intent of questions to keep the validity intact. Additional questions were added to collect contact information, the participant’s college of enrollment and degree program, and feedback about the survey. The HOLISTIC Cohort Study survey is presented in **Appendix 1** (tracked changes) and **Appendix 2** (clean copy).

### Adaptations of the HOLISTIC Cohort Study to increase feasibility during COVID-19 pandemic

The student research team and faculty mentors designed and implemented this study completely virtually throughout the duration of the research efforts. To improve feasibility for student participants to complete the survey during the COVID-19 pandemic, the team 1) obtained IRB-approved e-consent, rather than in-person consent; all participants received a copy of the signed and dated consent document via email; 2) the team relied on electronic communication to promote study participation and recruit students to complete the HOLISTIC Cohort Study survey. In-person recruitment (via the classroom, at research fairs, at campus events) was not feasible due to infection control precautions that were in-place at UIC. The team posted recruitment materials on-line and through e-mail, rather than using physical flyers. Information about the study was also made available on a dedicated webpage.^13^

### Data management

Study data were collected and managed using REDCap electronic data capture tools hosted at UIC.^12^ REDCap surveys from each respective health science college were exported into a Statistical Analysis System (SAS) dataset, with data labels and variable names provided for each question from the survey. Datasets were directly imported into SAS v9.4, and additional data cleaning was done to identify unusual responses and deal with missing data patterns. Upon completion of cleaning data, the study team re-checked all variables to make sure all responses were valid, and the data was stored with a data identifier to uniquely identify an object with an ID.

### Data analysis

All analyses were conducted using SAS v9.4. Response rate was defined as the number of participants who completed one or more questions in the survey and clicked the “submit” button on REDCap divided by the number of students invited to participate. Participants who did not click the “submit” button were considered non-responders. The Little’s Missing Completely at Random (MCAR) Test was used to test the null hypothesis that data for gender, race, and health science college was missing completely at random.^19^ Descriptive statistics were conducted using means and standard deviations for continuous variables, as well as using frequencies and percentages for categorical variables. Analyses were conducted among all participants and in participants stratified by their health science college.

## RESULTS

### Response rate

The Spring 2021 wave of recruitment resulted in 553 participants (10.8% of 5,118 eligible students; 27.7% of the 2,000-participant recruitment goal). Using Little’s MCAR Test on several key important variables (e.g., Gender, College enrolled, race, etc.) showed that these variables were missing completely at random. Each of these variables were tested for each health science college.

### Survey characteristics

The participants of the HOLISTIC Cohort Study (after the first of three recruitment waves) represent all seven UIC health science colleges and a wide variety of health science programs (**Table 4**). **Table 5** outlines the baseline characteristics of the first wave of participants. The average age of student participants is 27.3 years, 435 (78.8%) identify as female, and 137 (24.8%) identify as an underrepresented minority, defined as Hispanic, Black/African American, American Indian/Alaskan Native, and Multi-race. Overall, 465 (84.6%) participants reported being currently employed for wages. Just under one in five (94, 17%) participants reported “excellent” health. While 282 (51%) participants reported “0 days per month when their physical health is not good”, only 62 (11.2%) reported “0 days per month when their mental health is not good”. About 1 in 11 (52, 9.4%) reported having ever tested positive for COVID-19.

**Table 4:**
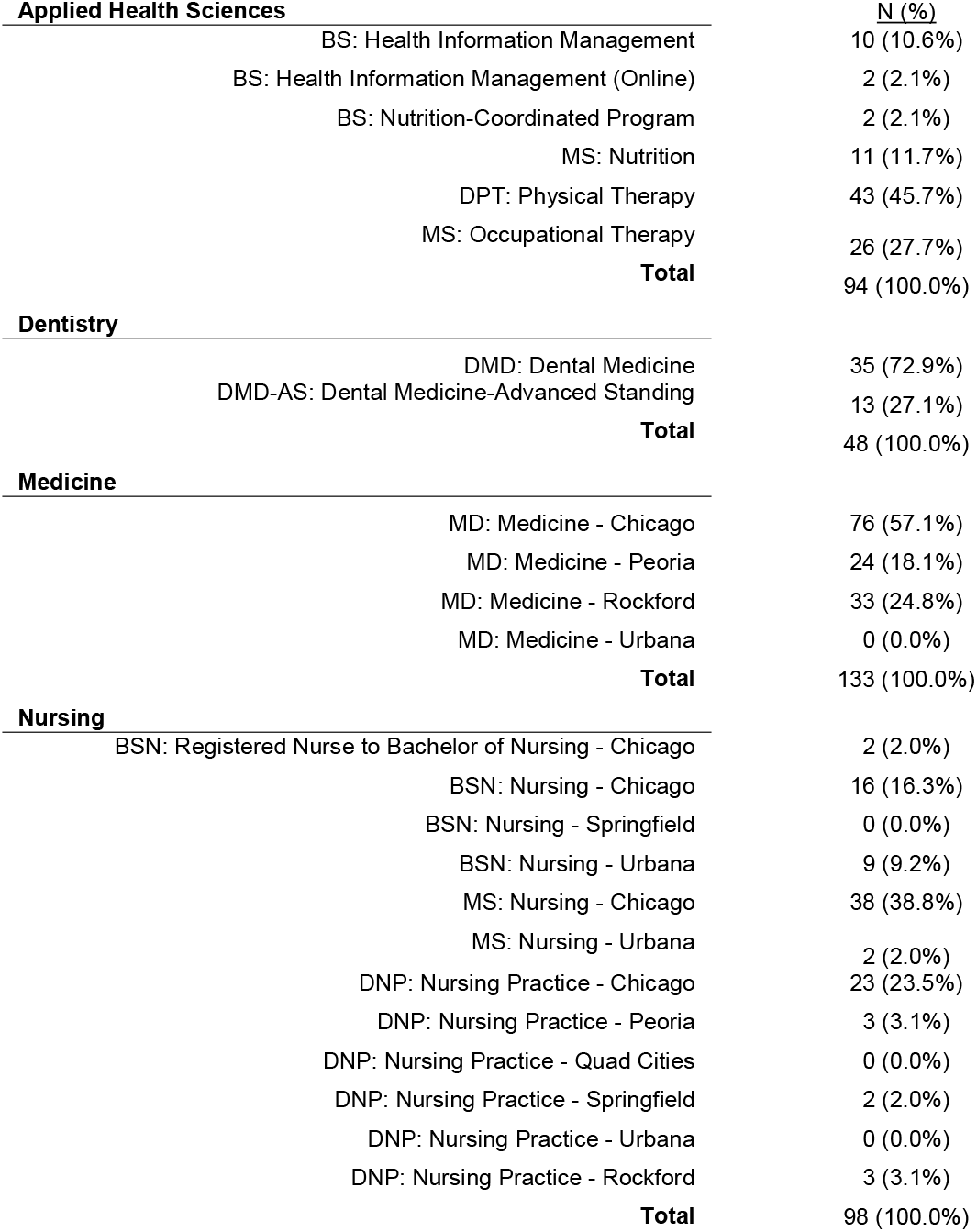
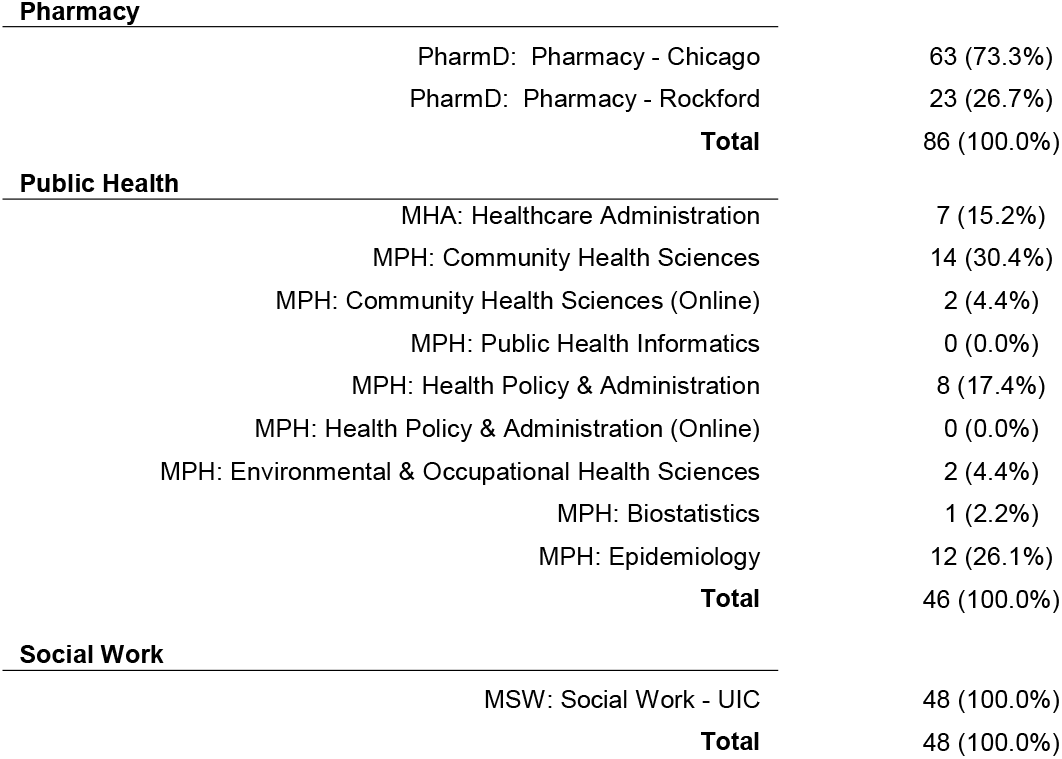
Degree programs of HOLISTIC cohort participants.

| **Applied Health Sciences** |  | <u>N (%)</u> |
| --- | --- | --- |
| BS: Health Information Management |  | 10 (10.6%) |
| BS: Health Information Management (Online) |  | 2 (2.1%) |
| BS: Nutrition-Coordinated Program |  | 2 (2.1%) |
| MS: Nutrition |  | 11 (11.7%) |
| DPT: Physical Therapy |  | 43 (45.7%) |
| MS: Occupational Therapy |  | 26 (27.7%) |
| **Total** |  | 94 (100.0%) |
| **Dentistry** |  |  |
| DMD: Dental Medicine |  | 35 (72.9%) |
| DMD-AS: Dental Medicine-Advanced Standing |  | 13 (27.1%) |
| **Total** |  | 48 (100.0%) |
| **Medicine** |  |  |
| MD: Medicine - Chicago |  | 76 (57.1%) |
| MD: Medicine - Peoria |  | 24 (18.1%) |
| MD: Medicine - Rockford |  | 33 (24.8%) |
| MD: Medicine - Urbana |  | 0 (0.0%) |
| **Total** |  | 133 (100.0%) |
| **Nursing** |  |  |
| BSN: Registered Nurse to Bachelor of Nursing - Chicago |  | 2 (2.0%) |
| BSN: Nursing - Chicago |  | 16 (16.3%) |
| BSN: Nursing - Springfield |  | 0 (0.0%) |
| BSN: Nursing - Urbana |  | 9 (9.2%) |
| MS: Nursing - Chicago |  | 38 (38.8%) |
| MS: Nursing - Urbana |  | 2 (2.0%) |
| DNP: Nursing Practice - Chicago |  | 23 (23.5%) |
| DNP: Nursing Practice - Peoria |  | 3 (3.1%) |
| DNP: Nursing Practice - Quad Cities |  | 0 (0.0%) |
| DNP: Nursing Practice - Springfield |  | 2 (2.0%) |
| DNP: Nursing Practice - Urbana |  | 0 (0.0%) |
| DNP: Nursing Practice - Rockford |  | 3 (3.1%) |
| **Total** |  | 98 (100.0%) |

**Pharmacy**
|  |  |
| --- | --- |
| PharmD: Pharmacy - Chicago | 63 (73.3%) |
| PharmD: Pharmacy - Rockford | 23 (26.7%) |
| **Total** | **86 (100.0%)** |

**Public Health**
|  |  |
| --- | --- |
| MHA: Healthcare Administration | 7 (15.2%) |
| MPH: Community Health Sciences | 14 (30.4%) |
| MPH: Community Health Sciences (Online) | 2 (4.4%) |
| MPH: Public Health Informatics | 0 (0.0%) |
| MPH: Health Policy & Administration | 8 (17.4%) |
| MPH: Health Policy & Administration (Online) | 0 (0.0%) |
| MPH: Environmental & Occupational Health Sciences | 2 (4.4%) |
| MPH: Biostatistics | 1 (2.2%) |
| MPH: Epidemiology | 12 (26.1%) |
| **Total** | **46 (100.0%)** |

**Social Work**
|  |  |
| --- | --- |
| MSW: Social Work - UIC | 48 (100.0%) |
| **Total** | **48 (100.0%)** |

**Table 5:** Baseline characteristics of the HOLISTIC cohort study participants.

| <u>Characteristic</u> | **Applied Health Sciences** | **Dentistry** | **Medicine** | **Nursing** | **Pharmacy** | **Public Health** | **Social Work** | **<u>Total</u>** |
| --- | --- | --- | --- | --- | --- | --- | --- | --- |
| **Total Observed Students Who Completed Survey** | 94 | 48 | 133 | 98 | 86 | 46 | 48 | 553 |
| **Age (Years): Mean ± SD** | 26.0 ± 3.2 | 29.2 ± 7.0 | 26.9 ± 2.3 | 28.4 ± 7.6 | 25.9 ± 4.2 | 27.2 ± 5.0 | 29.8 ± 6.8 | 27.3 ± 5.3 |
| **Gender: N (%)** |  |  |  |  |  |  |  |  |
| Male | 17 (18.1%) | 35 (74.5%) | 46 (34.6%) | 7 (7.1%) | 26 (30.2%) | 3 (6.5%) | 2 (4.2%) | 112 (20.3%) |
| Female | 77 (81.9%) | 11 (23.4%) | 85 (63.9%) | 90 (91.8%) | 59 (68.6%) | 43 (93.5%) | 46 (95.8%) | 435 (78.8%) |
| Other Response | 0 (0.0%) | 1 (2.1%) | 2 (1.5%) | 1 (1.0%) | 1 (1.2%) | 0 (0.0%) | 0 (0.0%) | 5 (0.9%) |
| **Ethnicity: N (%)** |  |  |  |  |  |  |  |  |
| Latino(a), Hispanic, or Spanish origin | 10 (10.6%) | 2 (4.2%) | 17 (12.8%) | 11 (11.2%) | 8 (9.3%) | 11 (23.9%) | 12 (25.0%) | 71 (12.8%) |
| Not Latino(a), Hispanic, or Spanish origin | 84 (89.4%) | 46 (95.8%) | 113 (85.0%) | 85 (86.7%) | 78 (90.7%) | 34 (73.9%) | 35 (72.9%) | 475 (85.9%) |
| Other Response | 0 (0.0%) | 0 (0.0%) | 3 (2.3%) | 2 (2.0%) | 0 (0.0%) | 1 (2.2%) | 1 (2.1%) | 7 (1.3%) |
| **Race: N (%)** |  |  |  |  |  |  |  |  |
| White | 57 (60.6%) | 18 (37.5%) | 73 (54.9%) | 72 (73.5%) | 26 (30.2%) | 30 (65.2%) | 29 (60.4%) | 305 (55.2%) |
| Black/African American | 1 (1.1%) | 4 (8.3%) | 6 (4.5%) | 3 (3.1%) | 5 (5.8%) | 4 (8.7%) | 3 (6.2%) | 26 (4.7%) |
| American Indian/Alaskan Native | 1 (1.1%) | 0 (0.0%) | 0 (0.0%) | 0 (0.0%) | 0 (0.0%) | 0 (0.0%) | 1 (2.1%) | 2 (0.4%) |
| Asian | 27 (28.7%) | 20 (41.7%) | 37 (27.8%) | 16 (16.3%) | 39 (45.3%) | 7 (15.2%) | 6 (12.5%) | 152 (27.5%) |
| Pacific Islander | 1 (1.1%) | 0 (0.0%) | 0 (0.0%) | 0 (0.0%) | 0 (0.0%) | 0 (0.0%) | 0 (0.0%) | 1 (0.2%) |
| Middle Eastern/North African | 0 (0.0%) | 1 (2.1%) | 1 (0.8%) | 0 (0.0%) | 0 (0.0%) | 0 (0.0%) | 0 (0.0%) | 2 (0.4%) |
| Multiracial | 6 (6.4%) | 4 (8.3%) | 14 (10.5%) | 6 (6.1%) | 14 (16.3%) | 5 (10.9%) | 7 (14.6%) | 56 (10.1%) |
| Other | 1 (1.1%) | 1 (2.1%) | 2 (1.5%) | 1 (1.0%) | 2 (2.3%) | 0 (0.0%) | 2 (4.2%) | 9 (1.6%) |
| **Underrepresented Minority\*: N (%)** |  |  |  |  |  |  |  |  |
| Yes | 16 (17.0%) | 10 (20.8%) | 33 (24.8%) | 19 (19.4%) | 24 (27.9%) | 16 (34.8%) | 19 (39.6%) | 137 (24.8%) |
| No | 78 (83.0%) | 38 (79.2%) | 100 (75.2%) | 79 (80.6%) | 62 (72.1%) | 30 (65.2%) | 29 (60.4%) | 416 (75.2%) |
| **Employment Status: N (%)** |  |  |  |  |  |  |  |  |
| Employed For Wages | 79 (84.0%) | 21 (45.7%) | 114 (85.7%) | 93 (94.9%) | 82 (96.5%) | 42 (91.3%) | 34 (70.8%) | 465 (84.6%) |
| Out of Work for >=1 Year | 5 (5.3%) | 25 (54.3%) | 8 (6.0%) | 1 (1.0%) | 1 (1.2%) | 4 (8.7%) | 12 (25.0%) | 56 (10.2%) |
| Out of Work for <1 Year | 10 (10.6%) | 0 (0.0%) | 11 (8.3%) | 3 (3.1%) | 2 (2.4%) | 0 (0.0%) | 1 (2.1%) | 27 (4.9%) |
| Unable to Work | 0 (0.0%) | 0 (0.0%) | 0 (0.0%) | 1 (1.0%) | 0 (0.0%) | 0 (0.0%) | 1 (2.1%) | 2 (0.4%) |
| **Health Status: N (%)** |  |  |  |  |  |  |  |  |
| Excellent | 19 (20.2%) | 11 (22.9%) | 28 (21.0%) | 9 (9.2%) | 11 (12.8%) | 11 (23.9%) | 5 (10.4%) | 94 (17.0%) |
| Very Good | 48 (51.1%) | 24 (50.0%) | 61 (45.9%) | 55 (56.1%) | 42 (48.8%) | 20 (43.5%) | 25 (52.1%) | 275 (49.7%) |
| Good | 26 (27.7%) | 10 (20.8%) | 37 (27.8%) | 32 (32.7%) | 27 (31.4%) | 11 (23.9%) | 16 (33.3%) | 159 (28.7%) |
| Fair | 1 (1.1%) | 3 (6.2%) | 6 (4.5%) | 2 (2.0%) | 5 (5.8%) | 3 (6.5%) | 2 (4.2%) | 22 (4.0%) |
| Poor | 0 (0.0%) | 0 (0.0%) | 1 (0.8%) | 0 (0.0%) | 1 (1.2%) | 1 (2.2%) | 0 (0.0%) | 3 (0.5%) |
| **Physical Health Last 30 Days: N (%)** |  |  |  |  |  |  |  |  |
| 0 Days Per Month When Physical Health Not Good | 56 (59.6%) | 30 (62.5%) | 68 (51.1%) | 37 (37.8%) | 44 (51.2%) | 19 (41.3%) | 28 (58.3%) | 282 (51.0%) |
| 1-13 Days Per Month When Physical Health Not Good | 34 (36.2%) | 16 (33.3%) | 59 (44.4%) | 54 (55.1%) | 36 (41.9%) | 23 (50.0%) | 18 (37.5%) | 240 (43.4%) |
| 14+ Days Per Month When Physical Health Not Good | 4 (4.3%) | 2 (4.2%) | 6 (4.5%) | 7 (7.1%) | 6 (7.0%) | 4 (8.7%) | 2 (4.2%) | 31 (5.6%) |
| **Mental Health Last 30 Days: N (%)** |  |  |  |  |  |  |  |  |
| 0 Days Per Month When Mental Health Not Good | 10 (10.6%) | 14 (29.2%) | 14 (10.5%) | 5 (5.1%) | 13 (15.1%) | 4 (8.7%) | 2 (4.2%) | 62 (11.2%) |
| 1-13 Days Per Month When Mental Health Not Good | 54 (57.5%) | 27 (56.3%) | 88 (66.2%) | 71 (72.5%) | 39 (45.4%) | 26 (56.5%) | 31 (64.6%) | 336 (60.8%) |
| 14+ Days Per Month When Mental Health Not Good | 30 (31.9%) | 7 (14.6%) | 31 (23.3%) | 22 (22.4%) | 34 (39.5%) | 16 (34.8%) | 15 (31.2%) | 155 (28.0%) |
| **Healthcare Access through Insurance: N (%)** |  |  |  |  |  |  |  |  |
| Yes | 85 (90.4%) | 43 (89.6%) | 125 (94.0%) | 90 (91.8%) | 78 (90.7%) | 43 (93.5%) | 46 (95.8%) | 510 (92.2%) |
| No | 6 (6.4%) | 5 (10.4%) | 5 (3.8%) | 5 (5.1%) | 5 (5.8%) | 3 (6.5%) | 2 (4.2%) | 31 (5.6%) |
| I don't know | 1 (1.1%) | 0 (0.0%) | 1 (0.8%) | 2 (2.0%) | 2 (2.3%) | 0 (0.0%) | 0 (0.0%) | 5 (0.9%) |
| Other Response | 2 (2.1%) | 0 (0.0%) | 2 (1.5%) | 1 (1.0%) | 1 (1.2%) | 0 (0.0%) | 0 (0.0%) | 7 (1.3%) |
| **COVID Test, Positive Result\*\*: N (%)** |  |  |  |  |  |  |  |  |
| Yes | 11 (11.7%) | 3 (6.4%) | 10 (7.5%) | 12 (12.2%) | 7 (8.1%) | 5 (10.9%) | 4 (8.3%) | 52 (9.4%) |
| No | 79 (84.0%) | 42 (89.4%) | 115 (86.5%) | 79 (80.6%) | 73 (84.9%) | 37 (80.4%) | 43 (89.6%) | 468 (84.8%) |
| Test wasn't able to provide results | 0 (0.0%) | 0 (0.0%) | 1 (0.8%) | 1 (1.0%) | 1 (1.2%) | 0 (0.0%) | 0 (0.0%) | 3 (0.5%) |
| Was Not Tested | 3 (3.2%) | 0 (0.0%) | 5 (3.8%) | 6 (6.1%) | 3 (3.5%) | 4 (8.7%) | 1 (2.1%) | 22 (4.0%) |
| Other Response | 1 (1.1%) | 2 (4.3%) | 2 (1.5%) | 0 (0.0%) | 2 (2.3%) | 0 (0.0%) | 0 (0.0%) | 7 (1.3%) |

\*Underrepresented minority defined as Hispanic, Black/African American, American Indian/Alaskan Native, or Multi-race
\*\*self-reported

## DISCUSSION

The HOLISTIC Cohort Study is designed to offer information about the physical, mental, and social health and occupational trajectory of students trained during the pandemic at an urban public university as they enter the health professional workforce over the next few years. After the first of three waves of recruitment, just over one-quarter (28%) of the 2,000-participant enrollment target has been enrolled. The important findings of the first wave of participants includes a strong female representation, which was partly expected due to the large female population of the overall health professional student population at UIC. The representation of underrepresented minorities in the first wave (24.8%) nearly matched the overall proportion of underrepresented minorities at UIC at large (26.7%). The data also reveals a high rate of student employment for wages, indicating that most health professional students are balancing course work with jobs. Next, most students consider themselves not to have excellent health with considerably more difficulty with mental health than physical health. While nearly all students have health care coverage, 31 (5.6%) students still have no healthcare coverage. This is alarming given UIC’s requirement for all health science students to have insurance as well as the availability of UIC’s “CampusCare” insurance. Nearly one in ten (9.4%) reported having ever had a positive test for COVID-19.

There are several unique aspects of the HOLISTIC Cohort Study. First, the study was designed and initiated during the COVID-19 pandemic. Next, the study was designed by an interprofessional student research team, providing the opportunity to support the professional and educational development of these students through interprofessional collaboration. Third, the design and implementation of the study including recruitment, enrollment, and completion of the survey was completed entirely online through virtual study team meetings, electronic approaches to participant recruitment, and use of e-consent due to infection control precautions during the COVID-19 pandemic. In addition, the HOLISTIC Cohort Study used questionnaires based on the BRFSS and WHO SAGE Working Group questionnaires, which allows all data collected throughout the course of the study to be compared with results from other studies and, in the case of the BRFSS, benchmarked with state and national trends in the U.S. Lastly, our study included study participants across seven UIC health science colleges, which provides a unique opportunity to compare the health, health-related behaviors, and occupational trajectories across the health professions trained during the COVID-19 pandemic.

### Challenges

During the first wave of recruitment, the primary challenge that was observed was a low response rate (about 11%). The team relied exclusively on emails to students and were unable to use in-person events or campaigns to raise awareness of the study due to infection control precautions. For the second and third waves of recruitment, the student research team will seek IRB approval to implement online newsletters, social media, class announcements, and virtual townhalls to improve visibility of the study. The team will also seek approval for in-person campaigns, such as the use of small-group information sessions about the HOLISTIC study and placing flyers on bulletin boards near classrooms or in cafeterias used by students. These approaches may need to be tailored to each health science college. When used in conjunction with invitations via email, we expect these virtual and in-person campaigns to further increase awareness about the HOLISTIC Cohort Study and, potentially, response rates.

## CONCLUSION

The HOLISTIC Cohort Study has established a platform for assessing the health of health professional students educated during a unique time in history - the COVID-19 pandemic. The longitudinal design will allow evaluation of changes in health and health behaviors that occur during the transition from student to working professionals. Preliminary results from participants enrolled in the first wave of recruitment demonstrate a cohort with diverse backgrounds and suggest the need for increased attention to supporting the health and wellbeing of health professional students.

## Supporting information

Appendix1-3_HOLISTIC Cohort Study

## Data Availability

All data produced in the present study are available upon reasonable request to the authors

## Acknowledgements

We would like to acknowledge the following individuals for their assistance in preparation of IRB materials, development of survey platform, and input on recruitment strategy: Ummesalmah Abdulbaseer, Ellie Braun, Julie DeLisa, Joann Huynh, Sai Illendula, Bansari Modhera, and Melissa Rutherfoord.

