## Appendix1-3_HOLISTIC Cohort Study for "Rationale and design of the Health Professional Students at the University of Illinois Chicago (HOLISTIC) Cohort Study"

**APPENDIX: HOLISTIC Cohort Study**

In this Appendix, we present a tracked-changes version of the HOLISTIC Cohort Study questionnaire (Appendix 1) and the final copy of the HOLISTIC Cohort Study questionnaire (Appendix 2). A demographics table of the eligible health professional students by college is also presented (Appendix 3).

**Appendix 1**

*Includes a track-changes version that describe changes to the original wording of the BRFSS and WHO SAGE Working Group questionnaires. Additions to the wording based on this feedback are presented with yellow highlights; deletions are presented with a crossed-out line over the word or sentence.*

**HOLISTIC QUESTIONNARIE**

The information you provide throughout this survey will help us understand the physical, emotional, and social health of students across our health sciences colleges. We ask that you answer each question as accurately as possible. The survey should take about 15 to 20 minutes to complete from start to finish. For any questions about why certain questions were included or to provide feedback, please contact the principal investigator at 312-413-2873.

**Contact Information** We would like your contact information to follow up with you in the future regarding this study as well as to email you a $5 Amazon e-gift card after completion of the questionnaire. You will receive the gift card to your primary email address within 2-3 business days of completing the survey. Staff in the Office of the Vice Chancellor for Health Affairs will keep your survey participation private and your responses will be kept confidential. The student leaders of the HOLISTIC study will not have access to individual student responses. We will present results of the study in a way that will not allow individual respondents to be identifiable.

Study ID (pre-populated)

1. What is your University ID number (UIN)? ________

1a. Please re-enter your University ID number (UIN): _________

2. What is your first name (required)?

3. What is your last name (required)?

4. What is your cell phone number (required)?

5. What is your primary email address (required)?

6. Do you have another email address (e.g., non-UIC gmail account)?

A. Yes

B. No

6a. [If Yes to #6] What is your other (secondary) email address?

7. Which UIC Health Science College are you enrolled in? *Only eligible programs are listed. If you do not see your degree program, you are not eligible for this study.*

A. UIC College of Applied Health Sciences
B. University of Illinois College of Medicine
C. UIC College of Dentistry
D. UIC College of Nursing
E. UIC College of Pharmacy
F. UIC School of Public Health
G. Jane Addams College of Social Work (Degree program: MSW:Social Work -UIC)

8. Which degree program are you enrolled in?

8a. *[If A is selected for #7]*

A. *BS: Health Information Management*
B. *BS: Health Information Management (Online)*
C. *BS: Nutrition-Coordinated Program*
D. *MS: Nutrition*
E. *DPT: Physical Therapy*
F. *MS: Occupational Therapy*

8b. *[If B is selected for #7]*

A. *MD: Medicine - Chicago*
B. *MD: Medicine - Peoria*
C. *MD: Medicine - Rockford*
D. *MD: Medicine -Urbana*

8c. *[If C is selected for #7]*

A. *DMD: Dental Medicine*
B. *DMD-AS: Dental Medicine-Advanced Standing*

8d. *[If D is selected for #7]*

A. *BSN: Registered Nurse to Bachelor of Nursing - Chicago*
B. *BSN: Nursing - Chicago*
C. *BSN: Nursing - Springfield*
D. *BSN: Nursing – Urbana*
E. *MS: Nursing - Chicago*
F. *MS: Nursing - Urbana*
G. *DNP: Nursing Practice -Chicago*
H. *DNP: Nursing Practice - Peoria*
I. *DNP: Nursing Practice – Quad Cities*
J. *DNP: Nursing Practice - Springfield*
K. *DNP: Nursing Practice – Urbana*
L. *DNP: Nursing Practice - Rockford*

8e. *[If E is selected for #7]*

A. *PharmD: Pharmacy – Chicago*
B. *PharmD: Pharmacy - Rockford*

8f*. [If F is selected for #7]*

A. *MHA: Healthcare Administration*
B. *MPH: Community Health Sciences (Online)*
C. *MPH: Community Health Sciences*
D. *MPH: Public Health Informatics*
E. *MPH: Health Policy & Administration (Online)*
F. *MPH: Health Policy & Administration*
G. *MPH: Environmental & Occupational Health Sciences*
H. *MPH: Biostatistics*
I. *MPH: Epidemiology*

**Core Section 1: Health Status**

9. Would you say that in general your health is

1. Excellent
2. Very Good
3. Good
4. Fair
5. Poor
6. ~~Don’t know/Not sure~~
7. ~~Refused~~

**Core Section 2: Healthy Days**

10. Now thinking about your physical health, which includes physical illness and injury, for how many days during the past 30 days was your physical health not good?

1. _ _ Number of days (0~~1~~ - 30)
2. ~~None~~
3. ~~Don’t know/Not sure~~
4. ~~Refused~~

11.  Now thinking about your mental health, which includes stress, depression, or difficulties with emotions, for how many days during the past 30 days was your mental health not good?

1. _ _ Number of days (0~~1~~ - 30)
2. ~~None~~
3. ~~Don’t know/Not sure~~
4. ~~Refused~~

12. During the past 30 days, for about how many days did poor physical or mental health keep you from doing your usual activities, such as self-care, work, or recreation? ~~(Do not ask this question and skip to next section if the previous 2 questions were none)~~

1. _ _ Number of days (0~~1~~ - 30)
2. ~~None~~
3. ~~Don’t know/Not sure~~
4. ~~Refused~~

**Core Section 3: Healthcare Access**

13. Do you have any kind of health care coverage, including health insurance, prepaid plans such as HMOs, or government plans such as Medicare or Indian Health Service?

1. Yes
2. No
3. ~~Don’t know/Not sure~~ I do not know
4. ~~Refused~~ Prefer not to answer

14. Do you have one person you think of as your personal doctor or health care provider?

1. Yes, only one
2. Yes, more than one
3. No ~~(If No, read: Is there more than one, or is there no person who you think of as your personal doctor or health care provider?)~~
4. ~~Don’t know/Not sure~~ I do not know
5. ~~Refused~~ Prefer not to answer

15. Was there a time in the past 12 months when you needed to see a doctor but could not because of cost?

1. Yes
2. No
3. ~~Don’t know/Not sure~~ I do not know
4. ~~Refused~~ Prefer not to answer

16. About how long has it been since you last visited a doctor for a routine checkup? ~~(Read if necessary: A routine checkup is a general physical exam, not an exam for a specific injury, illness, or condition.)~~

1. ~~Within the past year (Anytime less than 12 months ago)~~ Anytime less than or equal to one year ago
2. ~~Within the past 2 years (1 year but less than 2 years ago)~~ More than 1 year but less than or equal to 2 years ago
3. ~~Within the past 5 years (2 years but less than 5 years ago)~~ More than 2 years but less than 5 years ago
4. Five or more years ago
5. I do not know
6. Never
7. ~~Refused~~ Prefer not to answer

**Core Section 4: Hypertension Awareness**

17. Have you ever been told by a doctor, nurse, or other health professional that you have high blood pressure?

1. Yes
2. Yes, ~~but female told me~~ only during pregnancy
3. No
4. Told borderline high or prehypertensive
5. ~~Don’t know/Not sure~~ I do not know
6. ~~Refused~~ Prefer not to answer

18. [*If A or B to #17*] Are you currently taking prescription medicine for your high blood pressure?

1. Yes
2. No
3. ~~I do not know~~
4. ~~Refused~~

**Core Section 5: Cholesterol Awareness**

19. Blood cholesterol is a fatty substance found in the blood. About how long has it been since you last had your blood cholesterol checked? ~~(If 1 or 9, go to next section)~~

1. Never
2. ~~Within the past year (anytime less than one year ago)~~ Anytime less than or equal to one year ago
3. ~~Within the past 2 years (1 year but less than 2 years ago)~~ More than 1 year but less than or equal to 2 years ago
4. ~~Within the past 3 years (2 years but less than 3 years ago)~~ More than 2 years but less than or equal to 3 years ago
5. ~~Within the past 4 years (3 years but less than 4 years ago)~~ More than 3 years but less than or equal to 4 years ago
6. ~~Within the past 5 years (4 years but less than 5 years ago)~~ More than 4 years but less than 5 years ago
7. Five or more years ago
8. I do not know
9. Prefer not to answer

20. Have you ever been told by a doctor, nurse or other health professional that your blood cholesterol is high? ~~(If 2, 3 or 4 go to next section)~~

1. Yes
2. No
3. ~~Don’t know/Not sure~~ I do not know
4. ~~Refused~~ Prefer not to answer

21. [*If A to #20*] Are you currently taking medicine prescribed by your doctor or other health professional for your blood cholesterol?

1. Yes
2. No
3. ~~Don’t know/Not sure~~ I do not know
4. ~~Refused~~ Prefer not to answer

**Core Section 6: Chronic Health Conditions**

Has a doctor, nurse, or other health professional ever told you that you had any of the following:

22. Heart attack also called a myocardial infarction?

1. Yes
2. No
3. ~~Don’t know/Not sure~~ I do not know
4. ~~Refused~~ Prefer not to answer

23. Angina or coronary heart disease?

1. Yes
2. No
3. ~~Don’t know/Not sure~~ I do not know
4. ~~Refused~~ Prefer not to answer

24. A stroke?

1. Yes
2. No
3. ~~Don’t know/Not sure~~ I do not know
4. ~~Refused~~ Prefer not to answer

25. Asthma? *[If participant answers Yes, go to #26]*

1. Yes
2. No
3. ~~Don’t know/Not sure~~ I do not know
4. ~~Refused~~ Prefer not to answer

26. *[If Yes to #25]* Do you still have asthma?

1. Yes
2. No
3. ~~Don’t know/Not sure~~ I do not know
4. ~~Refused~~ Prefer not to answer

27. Skin cancer?

1. Yes
2. No
3. ~~Don’t know/Not sure~~ I do not know
4. ~~Refused~~ Prefer not to answer

28. Any other type of cancer?

1. Yes
2. No
3. ~~Don’t know/Not sure~~ I do not know
4. ~~Refused~~ Prefer not to answer

29. Chronic obstructive pulmonary disease (COPD), emphysema, or chronic bronchitis?

1. Yes
2. No
3. ~~Don’t know/Not sure~~ I do not know
4. ~~Refused~~ Prefer not to answer

30. Depressive disorder (including depression, major depression, dysthymia, or minor depression)?

1. Yes
2. No
3. ~~Don’t know/Not sure~~ I do not know
4. ~~Refused~~ Prefer not to answer

31. Kidney disease, *not* including kidney stones, bladder infection or incontinence? ~~(Incontinence is not being able to control urine flow.)~~

1. Yes
2. No
3. ~~Don’t know/Not sure~~ I do not know
4. ~~Refused~~ Prefer not to answer

32. Diabetes, *not* including diabetes during pregnancy, pre-diabetes, or borderline diabetes?

1. Yes
2. No
3. ~~Don’t know/Not sure~~ I do not know
4. ~~Refused~~ Prefer not to answer

~~C06.12 How old were you when you were told you had diabetes~~

1. ~~_ _ Code age in years [97 = 97 and older]~~
2. ~~Don‘t know / Not sure~~
3. ~~Refused~~

**~~Core Section 7: Arthritis~~**

**Core Section 8: Demographics**

33. What was your sex at birth? Was it male or female?

1. Male
2. Female
3. I do not know
4. Prefer not to answer

34. ~~What is your age?~~ What is your date of birth?

1. ~~_ _ Code age in years~~
2. ~~Don’t know / Not sure~~
3. ~~Refused~~

_ _/_ _/_ _ _ _

35. Are you Hispanic, Latino/a, or of Spanish origin?

1. Mexican, Mexican American, Chicano/a
2. Puerto Rican
3. Cuban
4. Another Hispanic, Latino/a, or Spanish origin
5. No
6. ~~Don’t know/Not sure~~ I do not know
7. ~~Refused~~ Prefer not to answer

36. Which one or more of the following would you say is your race? (*Select all that apply*)

1. White
2. Black or African American
3. American Indian or Alaska Native
4. Asian
   a. Asian Indian
   b. Chinese
   c. Filipino
   d. Japanese
   e. Korean
   f. Vietnamese
   g. Other Asian
5. Pacific Islander
   a. Native Hawaiian
   b. Guamanian or Chamorro
   c. Samoan
   d. Other Pacific Islander
6. Middle Eastern or North African
   a. Lebanese
   b. Iranian
   c. Egyptian
   d. Syrian
   e. Moroccan
   f. Algerian
   g. Other Middle Eastern or North African
7. Other
8. None of the above
9. ~~Don’t know/Not sure~~ I do not know
10. ~~Refused~~ Prefer not to answer

37. Which one of these groups would you say *best* represents your race? *(Select only one)*

1. White
2. Black or African American
3. American Indian or Alaska Native
4. Asian
   a. Asian Indian
   b. Chinese
   c. Filipino
   d. Japanese
   e. Korean
   f. Vietnamese
   g. Other Asian
5. Pacific Islander
   h. Native Hawaiian
   i. Guamanian or Chamorro
   j. Samoan
   k. Other Pacific Islander
6. Middle Eastern or North African
   a. Lebanese
   b. Iranian
   c. Egyptian
   d. Syrian
   e. Moroccan
   f. Algerian
   g. Other Middle Eastern or North African
7. Other
8. None of the above
9. ~~Don’t know/Not sure~~ I do not know
10. ~~Refused~~ Prefer not to answer

38. What is your marital status? (*Select one*)

1. Married
2. Divorced
3. Widowed
4. Separated
5. Never married/single
6. ~~A member of an unmarried couple~~ None of the above
7. ~~Refused~~ Prefer not to answer

39.   What is the highest grade or year of school you completed? *(Select one)*

1. Never attended school or only attended kindergarten
2. Grades 1 through 8 (Elementary)
3. Grades 9 through 11 (Some high school)
4. Grade 12 or GED (High school graduate)
5. College 1 year to 3 years (Some college or technical school)
6. College 4 years or more (College graduate)
7. Graduate degree (Masters, PhD, doctorate, etc.)
8. ~~Refused~~ Prefer not to answer

40. Do you own or rent your home? *(Select one)*

1. Own
2. Rent
3. Other arrangement ~~(Other arrangement may include group home, staying with friends or family without paying rent. Home is defined as the place where you live most of the time/the majority of the year. Read if necessary: We ask this question in order to compare health indicators among people with different housing situations.)~~
4. ~~Don’t know/Not sure~~ I do not know
5. ~~Refused~~ Prefer not to answer

41.   In what county do you currently live?

1. ~~_ _ _ANSI County Code~~ ______
2. ~~Don’t know / Not sure~~
3. ~~Refused~~

42. What is the ZIP Code where you currently live?

1. _ _ _ _ _
2. ~~Don’t know / Not sure~~
3. ~~Refused~~

43.   Not including cell phones or numbers used for computers, fax machines or security systems, do you have more than one telephone number (e.g. landline telephone) in your household?

1. Yes
2. No
3. ~~Don’t know/Not sure~~ I do not know
4. ~~Refused~~ Prefer not to answer

44. *[If A to #44]* How many of these telephone numbers are residential numbers?

1. Enter number (1-5) ____
2. Six or more
3. I do not know
4. None
5. Prefer not to answer

45. How many cell phones do you have for personal use? Include cell phones used for both business and personal use.

1. Enter number (1-5) _____
2. Six or more
3. ~~Don’t know/Not sure~~ I do not know
4. None
5. ~~Refused~~ Prefer not to answer

46. Have you ever served on active duty in the United States Armed Forces, either in the regular military or in a National Guard or military reserve unit? Active duty does not include training for the Reserves or National Guard, but DOES include activation, for example, for the Persian Gulf War.

1. Yes
2. No
3. ~~Don’t know/Not sure~~
4. ~~Refused~~ Prefer not to answer

47. Are you currently…*(Select all that apply)*

1. Employed for wages
2. Self-employed
3. Out of work for 1 year or more
4. Out of work for less than 1 year
5. A Homemaker
6. A Student
7. Retired
8. Unable to work
9. ~~Refused~~

48. How many children less than 18 years of age live in your household?

1. _ _ Number of Children
2. None
3. ~~Refused~~ Prefer not to answer

49. What is your annual household income from all sources?

1. Less than $10,000
2. $10,000 to $24,999
3. $25,000 to $34,999
4. $35,000 to $49,999
5. $50,000 to $74,999
6. $75,000 or more
7. I do not know
8. Prefer not to answer

50. About how much do you weigh without shoes? *(Select one)*

1. _ _ _ pounds
2. _ _ _ kilograms
3. ~~Don’t know/Not sure~~ I do not know
4. ~~Refused~~ Prefer not to answer

51. About how tall are you without shoes? *(Select one)*

1. _ _ feet _ _ inches
2. _ _ meters _ _centimeters
3. ~~Don’t know/Not sure~~ I do not know
4. ~~Refused~~ Prefer not to answer

52. *[If B to #34]* To your knowledge, are you now pregnant?

1. Yes
2. No
3. ~~Don’t know/Not sure~~ I do not know
4. ~~Refused~~ Prefer not to answer

53. Some people who are deaf or have serious difficulty hearing use assistive devices to communicate. Are you deaf or do you have serious difficulty hearing?

1. Yes
2. No
3. ~~Don’t know/Not sure~~ I do not know
4. ~~Refused~~ Prefer not to answer

54. Are you blind or do you have serious difficulty seeing, even when wearing glasses?

1. Yes
2. No
3. ~~Don’t know/Not sure~~ I do not know
4. ~~Refused~~ Prefer not to answer

55. Because of a physical, mental, or emotional condition, do you have serious difficulty concentrating, remembering, or making decisions?

1. Yes
2. No
3. ~~Don’t know/Not sure~~ I do not know
4. ~~Refused~~ Prefer not to answer

56. Do you have difficulty walking or climbing stairs?

1. Yes
2. No
3. ~~Don’t know/Not sure~~ I do not know
4. ~~Refused~~ Prefer not to answer

57. Do you have difficulty dressing or bathing?

1. Yes
2. No
3. ~~Don’t know/Not sure~~ I do not know
4. ~~Refused~~ Prefer not to answer

58. Because of a physical, mental, or emotional condition, do you have difficulty doing errands alone such as visiting a doctor’s office or shopping?

1. Yes
2. No
3. ~~Don’t know/Not sure~~ I do not know
4. ~~Refused~~ Prefer not to answer

**Core Section 9: Tobacco Use**

59. Have you smoked at least 100 cigarettes in your entire life? Do not include electronic cigarettes, herbal cigarettes, cigars, cigarillos, little cigars, pipes, bidis, kreteks, water pipes (hookahs) or marijuana, 5 packs = 100 cigarettes.

1. Yes
2. No
3. ~~Don’t know/Not sure~~ I do not know
4. ~~Refused~~ Prefer not to answer

60. Do you now smoke cigarettes every day, some days, or not at all?

1. Every day
2. Some days
3. Not at all
4. ~~Don’t know/Not sure~~ I do not know
5. ~~Refused~~ Prefer not to answer

61. During the past 12 months, have you stopped smoking for one day or longer because you were trying to quit smoking?

1. Yes
2. No
3. Not currently a smoker
4. ~~Don’t know/Not sure~~ I do not know
5. ~~Refused~~ Prefer not to answer

62. How long has it been since you last smoked a cigarette, even one or two puffs?

1. ~~Within the past month (less than 1 month ago)~~ Less than 1 month ago
1. ~~Within the past 3 months (More than 1 month but less than 3 months ago)~~ More than 1 month but less than or equal to 3 months ago
2. ~~Within the past 6 months (3months but less than 6 months ago)~~ More than 3 months but less than or equal to 6 months ago
3. ~~Within the past year (6 months but less than 1 year ago)~~ More than 6 months but less than or equal to 1 year ago
4. ~~Within the past 5 years (1 year but less than 5 years ago)~~ More than 1 year but less than or equal to 5 years ago
5. ~~Within the past 10 years (5 years but less than 10 years ago)~~ More than 5 years but less than 10 years ago
5. 10 years or more
6. Never smoked regularly
7. ~~Don’t know/Not sure~~ I do not know
8. ~~Refused~~ Prefer not to answer

63. Do you currently use chewing tobacco, snuff ~~(Read if necessary: Snus (Swedish for snuff) is a moist smokeless tobacco, usually sold in small pouches that are placed under the lip against the gum)~~, or snus every day, some days, or not at all?

1. Every day
2. Some days
3. Not at all
4. ~~Don’t know/Not sure~~ I do not know
5. ~~Refused~~ Prefer not to answer

**Core Section 10: Alcohol Consumption**

64. During the past 30 days, how many days per week or per month did you have at least one drink of any alcoholic beverage such as beer, wine, a malt beverage or liquor? (Select one)

1. _ _ Days per week
2. _ _ Days in past 30 days
3. No drinks in past 30 days
4. ~~Don’t know/Not sure~~ I do not know
5. ~~Refused~~ Prefer not to answer

65. One drink is equivalent to a 12-ounce beer, a 5-ounce glass of wine, or a drink with one shot of liquor. During the past 30 days, on the days when you drank, about how many drinks did you drink on the average? ~~(Read if necessary: A 40 ounce beer would count as 3 drinks, or a cocktail drink with 2 shots would count as 2 drinks.)~~

1. _ _ Number of drinks
2. ~~None.~~ No drinks in past 30 days
3. ~~Don’t know/Not sure~~ I do not know
4. ~~Refused~~ Prefer not to answer

66. Considering all types of alcoholic beverages, how many times during the past 30 days did you have X [If A for #34 then, X= 5; if B for #34, X=4; if C or D for #34, X=4] or more drinks on an occasion?

1. _ _ Number of times
2. None
3. ~~Don’t know/Not sure~~ I do not know
4. ~~Refused~~ Prefer not to answer

67. During the past 30 days, what is the largest number of drinks you had on any occasion?

1. _ _ Number of drinks
2. ~~Don’t know/Not sure~~ I do not know
3. ~~Refused~~ Prefer not to answer

**Core Section 11: Exercise (Physical Activity)**

68.During the past month, other than your regular job, did you participate in any physical activities or exercises such as ~~running, calisthenics, golf, gardening, or walking for exercise~~ running/walking, weightlifting, playing sports, or other physical activities?

1. Yes
2. No
3. ~~Don’t know/Not sure~~ I do not know
4. ~~Refused~~ Prefer not to answer

69. *[If A to #69]* Referring to the exercise list below, what type of physical activity or exercise did you spend the *most* time doing during the past month?

1. _ _ Specify from Physical Activity Coding List (below)
2. ~~Don’t know/Not sure~~ I do not know
3. ~~Refused~~ Prefer not to answer

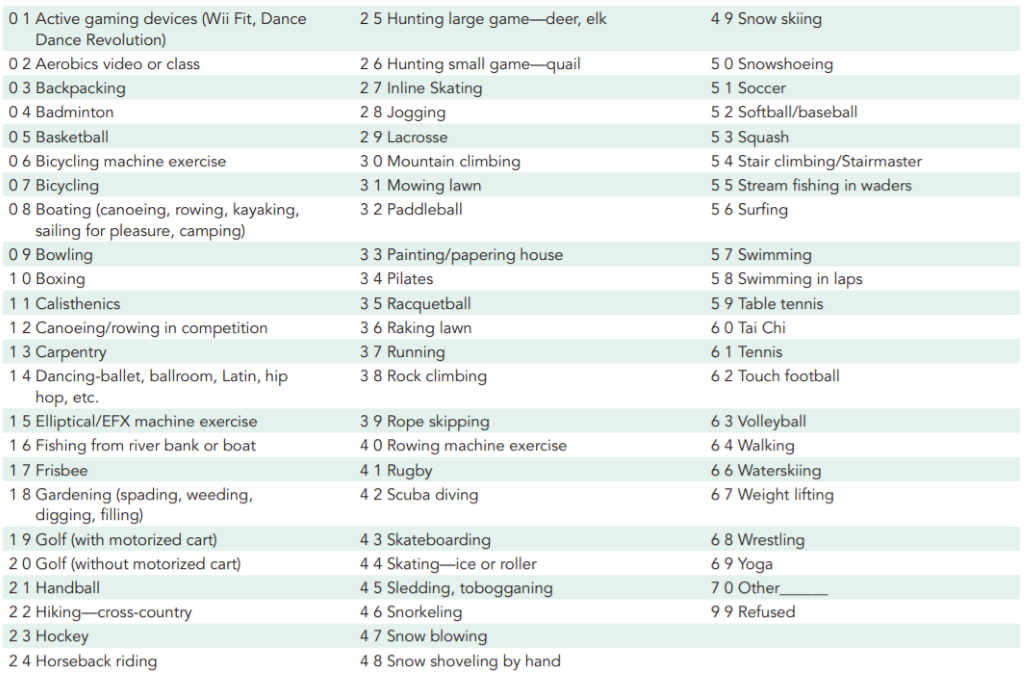

70. *[If A to #69]* How many times per week or per month did you take part in this activity during the past month?

1. _ _ Times per week
2. _ _ Times per month
3. ~~Don’t know/Not sure~~ I do not know
4. ~~Refused~~ Prefer not to answer

71. *[If A to #69]* And when you took part in this activity, for how many minutes did you usually keep at it?

1. ~~_:_ _Hours and minutes~~  _ _ Minutes
2. ~~Don’t know/Not sure~~ I do not know
3. ~~Refused~~ Prefer not to answer

72. *[If A to #69]* Referring to the exercise list, what other type of physical activity gave you the *next* most exercise during the past month?

1. _ _ _ Specify from physical activity list (below)
2. No other activity
3. ~~Don’t know/Not sure~~ I do not know
4. ~~Refused~~ Prefer not to answer

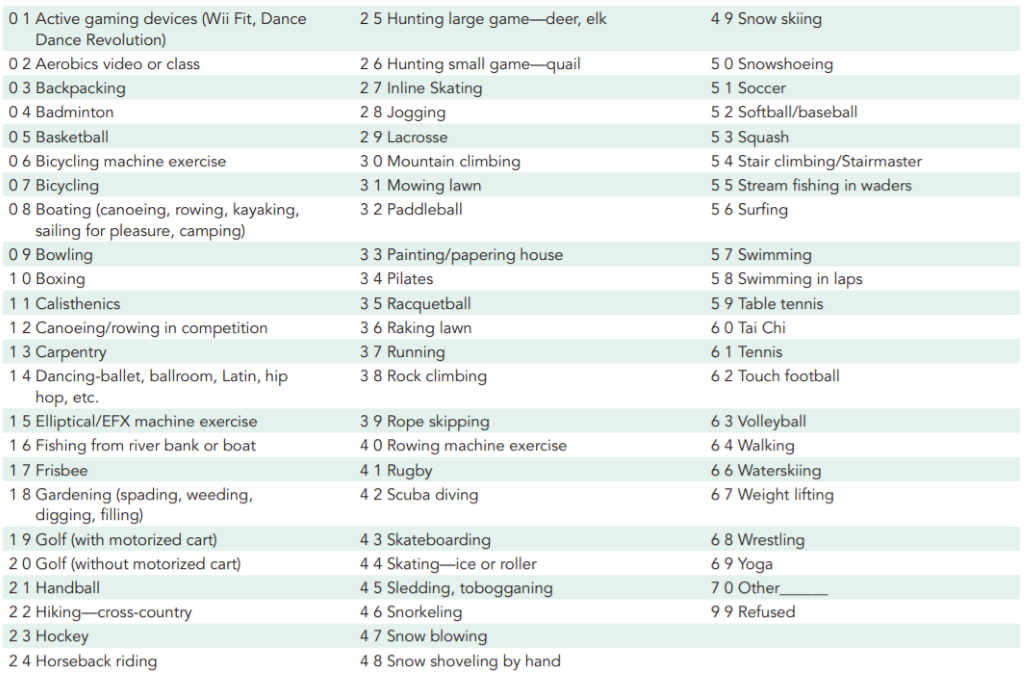

73. *[If A to #72]* How many times per week or per month did you take part in this activity during the past month?

1. _ _ Times per week
2. _ _ Times per month
3. ~~Don’t know/Not sure~~ I do not know
4. ~~Refused~~ Prefer not to answer

74. *[If A to #72]* And when you took part in this activity, for how many minutes did you usually keep at it?

1. ~~_:_ _Hours and minutes~~  _ _ Minutes
2. ~~Don’t know/Not sure~~ I do not know
3. ~~Refused~~ Prefer not to answer

75. *[If A to #69]* During the past month, how many times per week or per month did you do physical activities or exercises to strengthen your muscles? ~~(Do not count aerobic activities like walking, running, or bicycling. Count activities using your own body weight like yoga, sit-ups or push-ups and those using weight machines, free weights, or elastic bands.)~~

1. _ _ Times per week
2. _ _ Times per month
3. Never
4. ~~Don’t know/Not sure~~ I do not know
5. ~~Refused~~ Prefer not to answer

**Core Section 12: Fruits and Vegetables**

Now think about the foods you ate or drank during the past month, that is, the past 30 days, including meals and snacks.

76. Not including juices, how often did you eat fruit? ~~(If a respondent indicates that they consume a food item every day then enter the number of times per day. If the respondent indicates that they eat a food less than daily, then enter times per week or time per month. Do not enter time per day unless the respondent reports that he/she consumed that food item each day during the past month.Enter quantity in times per day, week, or month. If respondent gives a number without a time frame, ask “was that per day, week, or month?” Read if respondent asks what to include or says ‘I don’t know’: include fresh, frozen or canned fruit. Do not include dried fruits)~~

1. _ _ Day
2. _ _ Week
3. _ _ Month
4. Less than once a month
5. Never
6. ~~Don’t know/Not sure~~ I do not know
7. ~~Refused~~ Prefer not to answer

77. Not including fruit-flavored drinks or fruit juices with added sugar, how often did you drink 100% fruit juice such as apple or orange juice? Do not include fruit-flavored drinks with added sugar like cranberry cocktail, Hi-C, lemonade, KoolAid, Gatorade, Tampico, and sunny delight. Include only 100% pure juices or 100% juice blends.

1. _ _ Day
2. _ _ Week
3. _ _ Month
4. Less than once a month
5. Never
6. ~~Don’t know/Not sure~~ I do not know
7. ~~Refused~~ Prefer not to answer

78. How often did you eat a green leafy or lettuce salad, with or without other vegetables?

1. _ _ Day
2. _ _ Week
3. _ _ Month
4. Less than once a month
5. Never
6. ~~Don’t know/Not sure~~ I do not know
7. ~~Refused~~ Prefer not to answer

79. How often did you eat any kind of fried potatoes, including French fries, home fries, or hash browns?

1. _ _ Day
2. _ _ Week
3. _ _ Month
4. Less than once a month
5. Never
6. ~~Don’t know/Not sure~~ I do not know
7. ~~Refused~~ Prefer not to answer

80. How often did you eat any other kind of potatoes, or sweet potatoes, such as baked, boiled, mashed potatoes, or potato salad?

1. _ _ Day
2. _ _ Week
3. _ _ Month
4. Less than once a month
5. Never
6. ~~Don’t know/Not sure~~ I do not know
7. ~~Refused~~ Prefer not to answer

81. Not including lettuce salads and potatoes, how often did you eat other vegetables?

1. _ _ Day
2. _ _ Week
3. _ _ Month
4. Less than once a month
5. Never
6. ~~Don’t know/Not sure~~ I do not know
7. ~~Refused~~ Prefer not to answer

**Core Section 13: ~~Immunizations~~ Vaccinations**

82. During the past 12 months, have you had either a flu vaccine that was sprayed in your nose or a flu shot injected into your arm?

1. Yes
2. No
3. ~~Don’t know/Not sure~~ I do not know
4. ~~Refused~~ Prefer not to answer

83. *[If A to #83]* During what month and year did you receive your most recent flu vaccine that was sprayed in your nose or flu shot injected into your arm?

1. _ _ / _ _ _ _
2. ~~Don’t know/Not sure~~ I do not know
3. ~~Refused~~ Prefer not to answer

84. Have you received a tetanus shot in the past 10 years? ~~(If yes, ask: Was this Tdap, the tetanus shot that also has pertussis or whooping cough vaccine?)~~

1. Yes, received Tdap
2. Yes, received tetanus shot, but not Tdap
3. Yes, received tetanus shot but not sure what type
4. No, did not receive any tetanus shot in the past 10 years
5. ~~Don’t know/Not sure~~ I do not know
6. ~~Refused~~ Prefer not to answer

85. Have you ever had a pneumonia shot also known as a pneumococcal vaccine? ~~(Read if necessary: There are two types of pneumonia shots: polysaccharide, also known as Pneumovax, and conjugate, also known as Prevnar.)~~

1. Yes
2. No
3. ~~Don’t know/Not sure~~ I do not know
4. ~~Refused~~ Prefer not to answer

86. Have you ever had the Human Papilloma virus vaccination or HPV vaccination?

1. Yes
2. No
3. ~~Don’t know/Not sure~~ I do not know
4. ~~Refused~~ Prefer not to answer

87. *[If A to #87]* How many HPV shots did you receive?

1. _ _ Number of shots
2. All shots
3. ~~Don’t know/Not sure~~ I do not know
4. ~~Refused~~ Prefer not to answer

88. Have you had a positive test for COVID-19 or coronavirus?

1. Yes
2. No
3. Test wasn’t able to provide results
4. Was not tested
5. I do not know
6. Prefer not to answer

**Please let us know how much you agree or disagree with the statements below.**

89. A vaccine developed for COVID-19 or coronavirus would be important for my health.

1. Strongly Disagree
2. Disagree
3. Neutral
4. Agree
5. Strongly Agree

90. A vaccine developed for COVID-19 or coronavirus would be effective.

1. Strongly Disagree
2. Disagree
3. Neutral
4. Agree
5. Strongly Agree

91. Being vaccinated for COVID-19 or coronavirus would be important for the health of others in my community.

1. Strongly Disagree
2. Disagree
3. Neutral
4. Agree
5. Strongly Agree

92. All vaccines offered by the government program in my community are beneficial.

1. Strongly Disagree
2. Disagree
3. Neutral
4. Agree
5. Strongly Agree

93. New vaccines carry more risks than older vaccines.

1. Strongly Disagree
2. Disagree
3. Neutral
4. Agree
5. Strongly Agree

94. The information I receive about vaccines from public health officials is reliable and trustworthy. Public health officials include the Illinois Department of Public Health, Centers for Disease Control and Prevention (CDC), and the United States Food and Drug Administration (FDA).

1. Strongly Disagree
2. Disagree
3. Neutral
4. Agree
5. Strongly Agree

95. The information I receive about vaccines from my doctor or healthcare provider is reliable and trustworthy.

1. Strongly Disagree
2. Disagree
3. Neutral
4. Agree
5. Strongly Agree

96. Getting vaccines is a good way to protect me from disease.

1. Strongly Disagree
2. Disagree
3. Neutral
4. Agree
5. Strongly Agree

97. Generally, I do what my doctor or health care provider recommends about vaccines.

1. Strongly Disagree
2. Disagree
3. Neutral
4. Agree
5. Strongly Agree

98. I would be concerned about serious adverse effects of a vaccine developed for COVID-19 or coronavirus.

1. Strongly Disagree
2. Disagree
3. Neutral
4. Agree
5. Strongly Agree

99. I would receive a vaccine developed for COVID-19 or coronavirus.

1. Strongly Disagree
2. Disagree
3. Neutral
4. Agree
5. Strongly Agree

**~~Core section 14: HIV/AIDS~~**

**Module 1: Prediabetes**

100. Have you had a test for high blood sugar or diabetes within the past three years?

1. Yes
2. No
3. ~~Don’t know/Not sure~~ I do not know
4. ~~Refused~~

101. Have you ever been told by a doctor or other health professional that you have pre-diabetes or borderline diabetes?

1. Yes
2. Yes, during pregnancy
3. No
4. ~~Don’t know/Not sure~~ I do not know
5. ~~Refused~~

**Module 2: Diabetes**

102. *[If A to #32]* Are you now taking insulin?

1. Yes
2. No
3. ~~Don’t know/Not sure~~ I do not know
4. ~~Refused~~

103 *[If A to #32]* About how often do you check your blood for glucose or sugar? ~~(Read if necessary: Include times when checked by a family member or friend, but do not include times when checked by a health professional.Do not read: If the respondent uses a continuous glucose monitoring system (a sensor inserted under the skin to check glucose levels continuously),fill in ‘98 times per day.’)~~

1. _ _ Times per day
2. _ _ Times per week
3. _ _ Times per month
4. _ _ Times per year
5. Never
6. ~~Don’t know/Not sure~~ I do not know
7. ~~Refused~~

104 *[If A to #32]* Including times when checked by a family member or friend, about how often do you check your feet for any sores or irritations?

1. _ _ Times per day
2. _ _ Times per week
3. _ _ Times per month
4. _ _ Times per year
5. No feet
6. Never
7. ~~Don’t know/Not sure~~ I do not know
8. ~~Refused~~

105 *[If A to #32]* About how many times in the past 12 months have you seen a doctor, nurse, or other health professional for your diabetes?

1. _ _ Number of times
2. None
3. ~~Don’t know/Not sure~~ I do not know
4. ~~Refused~~

106 *[If A to #32]* About how many times in the past 12 months has a doctor, nurse, or other health professional checked your A1C? ~~(Read if necessary: A test for A-one-C measures the average level of blood sugar over the past three months)~~

1. _ _ Number of times
2. None
3. Never heard of A1C test
4. ~~Don’t know/Not sure~~ I do not know
5. ~~Refused~~

107 *[If A to #32]* About how many times in the past 12 months has a health professional checked your feet for any sores or irritations?

1. _ _ Number of times
2. None
3. ~~Don’t know/Not sure~~ I do not know
4. ~~Refused~~

108 *[If A to #32]* When was the last time you had an eye exam in which the pupils were dilated, making you temporarily sensitive to bright light?

1. Within the past month ~~(anytime less than 1 month ago)~~
2. ~~Within the past year (1 month but less than 12 months ago)~~ More than 1 month but less than or equal to 12 months ago
3. ~~Within the past 2 years (1 year but less than 2 years ago)~~ More than 1 year but less than 2 years ago
4. 2 or more years ago
5. ~~Don’t know/Not sure~~ I do not know
6. Never
7. ~~Refused~~

109. *[If A to #32]* Has a doctor ever told you that diabetes has affected your eyes or that you had retinopathy?

1. Yes
2. No
3. ~~Don’t know/Not sure~~ I do not know
4. ~~Refused~~

110. *[If A to #32]* Have you ever taken a course or class in how to manage your diabetes yourself?

1. Yes
2. No
3. ~~Don’t know/Not sure~~ I do not know
4. ~~Refused~~

**~~Module 3:ME/CFS~~**

**~~Module 4: Hepatitis Treatment~~**

**Module 5: HPV – Vaccinations  Core section 13 –Vaccinations**

**~~Module 6: Place of Flu Vaccination~~**

**~~Module 7: Shingles Vaccinations~~**

**~~Module 8: Lung Cancer Screening~~**

**Module 9: Breast and Cervical Cancer Screening**

111. [*If B to #34*] Have you ever had a mammogram? ~~(Interviewer Notes: A mammogram is an x-ray of each breast to look for breast cancer.)~~

1. Yes
2. No
3. ~~Don’t know/Not sure~~ I do not know
4. ~~Refused~~ Prefer not to answer

112. [*If A to #112*] How long has it been since you had your last mammogram?

1. Within the past year ~~(anytime less than 12 months ago)~~
2. ~~Within the past 2 years (1 year but less than 2 years ago)~~ More than 1 year but less than or equal to 2 years ago
3. ~~Within the past 3 years (2 years but less than 3 years ago)~~ More than 2 years but less than or equal to 3 years ago
4. ~~Within the past 5 years (3 years but less than 5 years ago)~~ More than 3 years but less than 5 years ago
5. 5 or more years ago
6. ~~Don’t know/Not sure~~ I do not know
7. ~~Refused~~ Prefer not to answer

113. *[If B to #34]* Have you ever had a Pap test?

1. Yes
2. No
3. ~~Don’t know/Not sure~~ I do not know
4. ~~Refused~~ Prefer not to answer

114.*[If A to #114]* How long has it been since you had your last Pap test?

Read if necessary:

1. Within the past year ~~(anytime less than 12 months ago)~~
2. ~~Within the past 2 years (1 year but less than 2 years ago)~~ More than 1 year but less than or equal to 2 years ago
3. ~~Within the past 3 years (2 years but less than 3years ago)~~ More than 2 years but less than or equal to 3 years ago
4. Within the past 5 years (3 years but less than 5 years ago) More than 3 years but less than 5 years ago
5. 5 or more years ago
6. ~~Don’t know/Not sure~~ I do not know
7. ~~Refused~~ Prefer not to answer

115. *[If A to #115]* An H.P.V. test is sometimes given with the Pap test for cervical cancer screening. Have you ever had an H.P.V. test?

1. Yes
2. No
3. ~~Don’t know/Not sure~~ I do not know
4. ~~Refused~~ Prefer not to answer

116. [*If A to #116*] How long has it been since you had your last H.P.V. test?

1. Within the past year ~~(anytime less than 12 months ago)~~
2. ~~Within the past 2 years (1 year but less than 2 years ago)~~ More than 1 year but less than or equal to 2 years ago
3. ~~Within the past 3 years (2 years but less than 3years ago)~~ More than 2 years but less than or equal to 3 years ago
4. Within the past 5 years (3 years but less than 5 years ago) More than 3 years but less than 5 years ago
5. 5 or more years ago
6. ~~Don’t know/Not sure~~ I do not know
7. ~~Refused~~ Prefer not to answer

117. [*If B to #34*] Have you had a hysterectomy? ~~(Read if necessary: A hysterectomy is an operation to remove the uterus (womb).)~~

1. Yes
2. No
3. ~~Don’t know/Not sure~~ I do not know
4. ~~Refused~~ Prefer not to answer

**~~Module 10: Prostate Cancer Screening~~**

**~~Module 11: Prostate Cancer Screening Decision Making~~**

**Module 12: Colorectal Cancer Screening**

118. A blood stool test is a test that may use a special kit at home to determine whether the stool contains blood. Have you ever had this test using a home kit?

1. Yes
2. No
3. ~~Don’t know/Not sure~~ I do not know
4. ~~Refused~~ Prefer not to answer

119. *[If A to #119]* How long has it been since you had your last blood stool test using a home kit?

1. Within the past year ~~(anytime less than 12 months ago)~~
2. ~~Within the past 2 years (1 year but less than 2 years ago)~~ More than 1 year but less than or equal to 2 years ago
3. ~~Within the past 3 years (2 years but less than 3years ago)~~ More than 2 years but less than or equal to 3 years ago
4. Within the past 5 years (3 years but less than 5 years ago) More than 3 years but less than 5 years ago
5. 5 or more years ago
6. ~~Don’t know/Not sure~~ I do not know
7. ~~Refused~~ Prefer not to answer

120. Sigmoidoscopy and colonoscopy are exams in which a tube is inserted into the rectum to view the colon for signs of cancer or other health problems. Have you ever had either of these exams?

1. Yes
2. No
3. ~~Don’t know/Not sure~~ I do not know
4. ~~Refused~~ Prefer not to answer

121. *[If A to #121]* For a sigmoidoscopy, a flexible tube is inserted into the rectum to look for problems. A colonoscopy is similar, but uses a longer tube, and you are usually given medication through a needle in your arm to make you sleepy and told to have someone else drive you home after the test. Was your most recent exam a sigmoidoscopy or a colonoscopy?

1. Sigmoidoscopy
2. Colonoscopy
3. ~~Don’t know/Not sure~~ I do not know
4. ~~Refused~~ Prefer not to answer

122. *[If A to #121]* How long has it been since you had your last sigmoidoscopy or colonoscopy?

1. Within the past year (anytime less than 12 months ago)
2. Within the past 2 years (1 year but less than 2 years ago)
3. Within the past 3 years (2 years but less than 3 years ago)
4. Within the past 5 years (3 years but less than 5 years ago)
5. Within the past 10 years (5 years but less than 10 years ago)
6. 10 or more years ago
7. ~~Don’t know/Not sure~~ I do not know
8. ~~Refused~~ Prefer not to answer

**~~Module 13: Cancer Survivorship~~**

**Module 14: Healthcare Access**

123. [*If A to #13*] What is the primary source of your health care coverage?

1. A plan purchased through an employer or union (including plans purchased through another person’s employer)
2. A plan that you or another family member buys ~~on your own~~
3. Medicare
4. Medicaid or other state program
5. TRICARE (formerly CAMPUS), VA, or Military
6. Alaska Native, Indian Health Service, Tribal Health Services
7. Some other source
8. None (no coverage)
9. ~~Don’t know/Not sure~~ I do not know
10. ~~Refused~~ Prefer not to answer

**~~Module 15: Aspirin for CVD Prevention~~**

**~~Module 16: Home/ Self-measured Blood Pressure~~**

**~~Module 17: Sodium or Salt-Related Behavior~~**

**~~Module 18: Indoor Tanning~~**

**~~Module 19: Excess Sun Exposure~~**

**~~Module 20: Cognitive Decline~~**

**~~Module 21: Caregiver~~**

**Module 22: Adverse Childhood Experiences.**

~~I'd like to ask you some questions about events that happened during your childhood. This information will allow us to better understand problems that may occur early in life, and may help others in the future. This is a sensitive topic and some people may feel uncomfortable with these questions. At the end of this section, I will give you a phone number for an organization that can provide information and referral for these issues. Please keep in mind that you can ask me to skip any question you do not want to answer. All questions refer to the time period before you were 18 years of age.~~

The following section will ask sensitive questions about depression, substance abuse, domestic violence, and sexual abuse within your family unit. We acknowledge that these questions may be difficult to answer and understand if certain questions will be answered with “Prefer not to answer”. We would also like to let you know that if you would like resources for mental health, substance abuse, domestic violence, and sexual abuse, please contact the UIC Counseling Center at (312) 996-3490 or visit <https://counseling.uic.edu/>

124. Now, looking back before you were 18 years of age, did you live with anyone who was depressed, mentally ill, or suicidal?

1. Yes
2. No
3. ~~Don’t know/Not sure~~ I do not know
4. ~~Refused~~ Prefer not to answer

125. Did you live with anyone who was a problem drinker or alcoholic?

1. Yes
2. No
3. ~~Don’t know/Not sure~~ I do not know
4. ~~Refused~~ Prefer not to answer

126. Did you live with anyone who used illegal street drugs or who abused prescription medications?

1. Yes
2. No
3. ~~Don’t know/Not sure~~ I do not know
4. ~~Refused~~ Prefer not to answer

127. Did you live with anyone who served time or was sentenced to serve time in a prison, jail, or other correctional facility?

1. Yes
2. No
3. ~~Don’t know/Not sure~~ I do not know
4. ~~Refused~~ Prefer not to answer

128. Were your parents separated or divorced?

1. Yes
2. ~~Parents not married.~~ No, parents still married
3. Parents never married
4. ~~Don’t know/Not sure~~ I do not know
5. ~~Refused~~ Prefer not to answer

129. How often did your parents or adults in your home ever slap, hit, kick, punch, or beat each other up?

1. Never
2. Once
3. More than once
4. ~~Don’t know/Not sure~~ I do not know
5. ~~Refused~~ Prefer not to answer

130. Not including spanking (before age 18), how often did a parent or adult in your home ever hit, beat, kick, or physically hurt you in any way?

1. Never
2. Once
3. More than once
4. ~~Don’t know/Not sure~~ I do not know
5. ~~Refused~~ Prefer not to answer

131. How often did a parent or adult in your home ever swear at you, insult you, or put you down?

1. Never
2. Once
3. More than once
4. ~~Don’t know/Not sure~~ I do not know
5. ~~Refused~~ Prefer not to answer

132. ~~How often did anyone at least 5 years older than you or an adult, ever touch you sexually?~~ How often did anyone ever touch you sexually without consent?

1. Never
2. Once
3. More than once
4. ~~Don’t know/Not sure~~ I do not know
5. ~~Refused~~ Prefer not to answer

133. ~~How often did anyone at least 5 years older than you or an adult, try to make you touch them sexually?~~ How often did anyone try to make you touch them sexually?

1. Never
2. Once
3. More than once
4. ~~Don’t know/Not sure~~ I do not know
5. ~~Refused~~ Prefer not to answer

134. ~~How often did anyone at least 5 years older than you or an adult, force you to have sex?~~ How often did anyone force you to have sex?

1. Never
2. Once
3. More than once
4. ~~Don’t know/Not sure~~ I do not know
5. ~~Refused~~ Prefer not to answer

**~~Module 23: Family Planning~~**

**~~Module 24: Alcohol Screening & Brief Intervention (ASBI)~~**

**Module 25: Marijuana Use**

135. During the past 30 days, on how many occasions did you use marijuana or cannabis? ~~(Marijuana and cannabis include both CBD and THC products.)~~

1. _ _ Number of days
2. None
3. ~~Don’t know/Not sure~~ I do not know
4. ~~Refused~~ Prefer not to answer

136. *[If A to #136]* During the past 30 days, which one of the following ways did you use marijuana the most often?

1. Smoke it (for example, in a joint, bong, pipe, or blunt)
2. Eat it (for example, in brownies, cakes, cookies, or candy)
3. Drink it (for example, in tea, cola, or alcohol)
4. Vaporize it (for example, in an e-cigarette-like vaporizer or another vaporizing device)
5. Dab it (for example, using waxes or concentrates)
6. Use it some other way
7. ~~Don’t know/Not sure~~ I do not know
8. ~~Refused~~ Prefer not to answer

137. *[If A to #136]* When you used marijuana or cannabis during the past 30 days, was it usually

1. For medical reasons (to treat or decrease symptoms of a health condition)
2. For non-medical reasons (like to have fun or fit in)
3. For both medical and non-medical reasons
4. ~~Don’t know/Not sure~~ I do not know
5. ~~Refused~~ Prefer not to answer

**~~Module 26: Industry and Occupation~~**

**~~Module 27: Food Stamps~~**

**Module 28: Sex at Birth  To our Core Section 8: Demographics**

**Module 29: Sexual Orientation and Gender Identity**

138. Which of the following best represents how you think of yourself? ~~(Interviewer Notes: Read if necessary: We ask this question in orderto better understand the health and health care needs of people with different sexual orientations.)~~

1. Gay
2. Straight, that is not gay
3. Bisexual
4. Something else
5. ~~I don't know the answer/ The respondent did not understand the~~ question I do not know
6. ~~Refused~~ Prefer not to answer

139 Do you consider yourself to be transgender? ~~(Interview Notes: Read if necessary: Some people describe themselves as transgender when they experience a different gender identity from their sex at birth. For example, a person born into a male body, but who feels female or lives as a woman would be transgender. Some transgender people change their physical appearance so that it matches their internal gender identity. Some transgender people take hormones and some have surgery. A transgender person may be of any sexual orientation –straight, gay, lesbian, or bisexual. If asked about definition of gender non-conforming: Some people think of themselves as gender non-conforming when they do not identify only as a man or only as a woman.)~~

1. Yes, transgender, male to female
2. Yes, transgender, female to male
3. Yes, transgender, gender nonconforming
4. No
5. ~~Don’t know/Not sure~~ I do not know
6. ~~Refused~~ Prefer not to answer

**~~Module 30: Random Child Selection~~**

**~~Module 31: Childhood Asthma Prevalence~~**

**Feedback**

140. How was your experience completing the HOLISTIC survey?

1. Very good
2. Good
3. Neutral
4. Poor
5. Very poor

141. Do you have any feedback about the HOLISTIC survey?

1. ____________

This is the end of our questionnaire! If you have any concerns about your physical or mental health, UIC has a variety of places to get assistance. One place to access assistance is the U and I Care program, which is an initiative within the UIC Student Affairs Office that helps connect members of the campus community to many different services and programs. Your college’s Office of Student Affairs is also a place to seek assistance.

You will receive a link to an Amazon.com electronic gift card for $5 to your primary email address within the next 10 days.

Please feel free to contact the study team at 312-413-2873 or at if you have any questions.

**Thank you for your participation!**

**Appendix 2**

*Includes the final, clean-copy version of the survey used in the HOLISTIC Cohort Study.*

**HOLISTIC QUESTIONNARIE**

The following questions will explore your health behaviors, access to care, past medical diagnoses, and demographic characteristics. The survey should take about 15 to 20 minutes to complete from start to finish. For any questions about why certain questions were included or to provide feedback, please contact the principal investigator at 312-413-2873.

**Contact Information** We would like your contact information to follow up with you in the future regarding this study as well as to email you a $5 Amazon e-gift card after completion of the questionnaire. You will receive the gift card to your primary email address within 1-2 business days of completing the survey. Your contact information will only be used for study purposes by study staff, and the study results will be coded without contact information before data analysis is conducted.

1. Study ID: (pre-populated)

2. What is your first name (required)?

3. What is your last name (required)?

4. What is your cell phone number (required)?

5. What is your primary email address (required)?

6. Do you have another email address (e.g., non-UIC gmail account)?

A. Yes

B. No

6a. [If Yes to #6] What is your other (secondary) email address?

7. Which UIC Health Science College are you enrolled in? *Only eligible programs are listed. If you do not see your degree program, you are not eligible for this study.*

H. UIC College of Applied Health Sciences
I. University of Illinois College of Medicine
J. UIC College of Dentistry
K. UIC College of Nursing
L. UIC College of Pharmacy
M. UIC School of Public Health
N. Jane Addams College of Social Work (Degree program: MSW:Social Work -UIC)

8. Which degree program are you enrolled in?

8a. *[If A is selected for #7]*

G. *BS: Health Information Management*
H. *BS: Health Information Management (Online)*
I. *BS: Nutrition-Coordinated Program*
J. *MS: Nutrition*
K. *DPT: Physical Therapy*
L. *MS: Occupational Therapy*

8b. *[If B is selected for #7]*

E. *MD: Medicine - Chicago*
F. *MD: Medicine - Peoria*
G. *MD: Medicine - Rockford*
H. *MD: Medicine -Urbana*

8c. *[If C is selected for #7]*

C. *DMD: Dental Medicine*
D. *DMD-AS: Dental Medicine-Advanced Standing*

8d. *[If D is selected for #7]*

M. *BSN: Registered Nurse to Bachelor of Nursing - Chicago*
N. *BSN: Nursing - Chicago*
O. *BSN: Nursing - Springfield*
P. *BSN: Nursing – Urbana*
Q. *MS: Nursing - Chicago*
R. *MS: Nursing - Urbana*
S. *DNP: Nursing Practice -Chicago*
T. *DNP: Nursing Practice - Peoria*
U. *DNP: Nursing Practice – Quad Cities*
V. *DNP: Nursing Practice - Springfield*
W. *DNP: Nursing Practice – Urbana*
X. *DNP: Nursing Practice - Rockford*

8e. *[If E is selected for #7]*

C. *PharmD: Pharmacy – Chicago*
D. *PharmD: Pharmacy - Rockford*

8f*. [If F is selected for #7]*

J. *MHA: Healthcare Administration*
K. *MPH: Community Health Sciences (Online)*
L. *MPH: Community Health Sciences*
M. *MPH: Public Health Informatics*
N. *MPH: Health Policy & Administration (Online)*
O. *MPH: Health Policy & Administration*
P. *MPH: Environmental & Occupational Health Sciences*
Q. *MPH: Biostatistics*
R. *MPH: Epidemiology*

**Core Section 1: Health Status**

9. Would you say that in general your health is

8. Excellent
9. Very Good
10. Good
11. Fair
12. Poor

**Core Section 2: Healthy Days**

10. Now thinking about your physical health, which includes physical illness and injury, for how many days during the past 30 days was your physical health not good?

_ _ Number of days (0-30)

11.  Now thinking about your mental health, which includes stress, depression, or difficulties with emotions, for how many days during the past 30 days was your mental health not good?

_ _ Number of days (0-30)

12. During the past 30 days, for about how many days did poor physical or mental health keep you from doing your usual activities, such as self-care, work, or recreation?

_ _ Number of days (0-30)

**Core Section 3: Healthcare Access**

13. Do you have any kind of health care coverage, including health insurance, prepaid plans such as HMOs, or government plans such as Medicare or Indian Health Service?

5. Yes
6. No
7. I do not know
8. Prefer not to answer

14. Do you have one person you think of as your personal doctor or health care provider?

6. Yes, only one
7. Yes, more than one
8. No
9. I do not know
10. Prefer not to answer

15. Was there a time in the past 12 months when you needed to see a doctor but could not because of cost?

5. Yes
6. No
7. I do not know
8. Prefer not to answer

16. About how long has it been since you last visited a doctor for a routine checkup?

8. Anytime less than or equal to one year ago
9. More than 1 year but less than or equal to 2 years ago
10. More than 2 years but less than 5 years ago
11. Five or more years ago
12. I do not know
13. Never
14. Prefer not to answer

**Core Section 4: Hypertension Awareness**

17. Have you ever been told by a doctor, nurse, or other health professional that you have high blood pressure?

7. Yes
8. Yes, only during pregnancy
9. No
10. Told borderline high or prehypertensive
11. I do not know
12. Prefer not to answer

18. [*If A or B to #17*] Are you currently taking prescription medicine for your high blood pressure?

5. Yes
6. No

**Core Section 5: Cholesterol Awareness**

19. Blood cholesterol is a fatty substance found in the blood. About how long has it been since you last had your blood cholesterol checked?

10. Never
11. Anytime less than or equal to one year ago
12. More than 1 year but less than or equal to 2 years ago
13. More than 2 years but less than or equal to 3 years ago
14. More than 3 years but less than or equal to 4 years ago
15. More than 4 years but less than 5 years ago
16. Five or more years ago
17. I do not know
18. Prefer not to answer

20. Have you ever been told by a doctor, nurse or other health professional that your blood cholesterol is high?

5. Yes
6. No
7. I do not know
8. Prefer not to answer

21. [*If A to #20*] Are you currently taking medicine prescribed by your doctor or other health professional for your blood cholesterol?

5. Yes
6. No
7. I do not know
8. Prefer not to answer

**Core Section 6: Chronic Health Conditions**

Has a doctor, nurse, or other health professional ever told you that you had any of the following:

22. Heart attack also called a myocardial infarction?

5. Yes
6. No
7. I do not know
8. Prefer not to answer

23. Angina or coronary heart disease?

5. Yes
6. No
7. I do not know
8. Prefer not to answer

24. A stroke?

5. Yes
6. No
7. I do not know
8. Prefer not to answer

25. Asthma? *[If participant answers Yes, go to #26]*

5. Yes
6. No
7. I do not know
8. Prefer not to answer

26. *[If Yes to #25]* Do you still have asthma?

5. Yes
6. No
7. I do not know
8. Prefer not to answer

27. Skin cancer?

5. Yes
6. No
7. I do not know
8. Prefer not to answer

28. Any other type of cancer?

5. Yes
6. No
7. I do not know
8. Prefer not to answer

29. Chronic obstructive pulmonary disease (COPD), emphysema, or chronic bronchitis?

5. Yes
6. No
7. I do not know
8. Prefer not to answer

30. Depressive disorder (including depression, major depression, dysthymia, or minor depression)?

5. Yes
6. No
7. I do not know
8. Prefer not to answer

31. Kidney disease, *not* including kidney stones, bladder infection or incontinence? (Incontinence is not being able to control urine flow.)

5. Yes
6. No
7. I do not know
8. Prefer not to answer

32. Diabetes, *not* including diabetes during pregnancy, pre-diabetes, or borderline diabetes?

5. Yes
6. No
7. I do not know
8. Prefer not to answer

**Core Section 8: Demographics**

33. What was your sex at birth? Was it male or female?

5. Male
6. Female
7. I do not know
8. Prefer not to answer

34. What is your date of birth?

_ _/_ _/_ _ _ _

35. Are you Hispanic, Latino/a, or of Spanish origin?

8. Mexican, Mexican American, Chicano/a
9. Puerto Rican
10. Cuban
11. Another Hispanic, Latino/a, or Spanish origin
12. No
13. I do not know
14. Prefer not to answer

36. Which one or more of the following would you say is your race? (*Select all that apply*)

11. White
12. Black or African American
13. American Indian or Alaska Native
14. Asian
    a. Asian Indian
    b. Chinese
    c. Filipino
    d. Japanese
    e. Korean
    f. Vietnamese
    g. Other Asian
15. Pacific Islander
    a. Native Hawaiian
    b. Guamanian or Chamorro
    c. Samoan
    d. Other Pacific Islander
16. Middle Eastern or North African
    a. Lebanese
    b. Iranian
    c. Egyptian
    d. Syrian
    e. Moroccan
    f. Algerian
    g. Other Middle Eastern or North African
17. Other
18. None of the above
19. I do not know
20. Prefer not to answer

37. Which one of these groups would you say *best* represents your race? *(Select only one)*

11. White
12. Black or African American
13. American Indian or Alaska Native
14. Asian
    l. Asian Indian
    m. Chinese
    n. Filipino
    o. Japanese
    p. Korean
    q. Vietnamese
    r. Other Asian
15. Pacific Islander
    s. Native Hawaiian
    t. Guamanian or Chamorro
    u. Samoan
    v. Other Pacific Islander
16. Middle Eastern or North African
    a. Lebanese
    b. Iranian
    c. Egyptian
    d. Syrian
    e. Moroccan
    f. Algerian
    g. Other Middle Eastern or North African
17. Other
18. None of the above
19. I do not know
20. Prefer not to answer

38. What is your marital status? (*Select one*)

8. Married
9. Divorced
10. Widowed
11. Separated
12. Never married/single
14. None of the abovePrefer not to answer

39.   What is the highest grade or year of school you completed? *(Select one)*

9. Never attended school or only attended kindergarten
10. Grades 1 through 8 (Elementary)
11. Grades 9 through 11 (Some high school)
12. Grade 12 or GED (High school graduate)
13. College 1 year to 3 years (Some college or technical school)
14. College 4 years or more (College graduate)
15. Graduate degree (Masters, PhD, doctorate, etc.)
16. Prefer not to answer

40. Do you own or rent your home? *(Select one)*

6. Own
7. Rent
8. Other arrangement
9. I do not know
10. Prefer not to answer

41.   In what county do you currently live?

42. What is the ZIP Code where you currently live?

43.   Not including cell phones or numbers used for computers, fax machines or security systems, do you have more than one telephone number (e.g. landline telephone) in your household?

5. Yes
6. No
7. I do not know
8. Prefer not to answer

44. *[If A to #44]* How many of these telephone numbers are residential numbers?

6. Enter number (1-5) ____
7. Six or more
8. I do not know
9. None
10. Prefer not to answer

45. How many cell phones do you have for personal use? Include cell phones used for both business and personal use.

6. Enter number (1-5) _____
7. Six or more
8. I do not know
9. None
10. Prefer not to answer

46. Have you ever served on active duty in the United States Armed Forces, either in the regular military or in a National Guard or military reserve unit? Active duty does not include training for the Reserves or National Guard, but DOES include activation, for example, for the Persian Gulf War.

5. Yes
6. No
7. Prefer not to answer

47. Are you currently…*(Select all that apply)*

10. Employed for wages
11. Self-employed
12. Out of work for 1 year or more
13. Out of work for less than 1 year
14. A Homemaker
15. A Student
16. Retired
17. Unable to work

48. How many children less than 18 years of age live in your household?

4. _ _ Number of Children
5. None
6. Prefer not to answer

49. What is your annual household income from all sources?

9. Less than $10,000
10. $10,000 to $24,999
11. $25,000 to $34,999
12. $35,000 to $49,999
13. $50,000 to $74,999
14. $75,000 or more
15. I do not know
16. Prefer not to answer

50. About how much do you weigh without shoes? *(Select one)*

5. _ _ _ pounds
6. _ _ _ kilograms
7. I do not know
8. Prefer not to answer

51. About how tall are you without shoes? *(Select one)*

5. _ _ feet _ _ inches
6. _ _ meters _ _centimeters
7. I do not know
8. Prefer not to answer

52. *[If B to #34]* To your knowledge, are you now pregnant?

5. Yes
6. No
7. I do not know
8. Prefer not to answer

53. Some people who are deaf or have serious difficulty hearing use assistive devices to communicate. Are you deaf or do you have serious difficulty hearing?

5. Yes
6. No
7. I do not know
8. Prefer not to answer

54. Are you blind or do you have serious difficulty seeing, even when wearing glasses?

5. Yes
6. No
7. I do not know
8. Prefer not to answer

55. Because of a physical, mental, or emotional condition, do you have serious difficulty concentrating, remembering, or making decisions?

5. Yes
6. No
7. I do not know
8. Prefer not to answer

56. Do you have difficulty walking or climbing stairs?

5. Yes
6. No
7. I do not know
8. Prefer not to answer

57. Do you have difficulty dressing or bathing?

5. Yes
6. No
7. I do not know
8. Prefer not to answer

58. Because of a physical, mental, or emotional condition, do you have difficulty doing errands alone such as visiting a doctor’s office or shopping?

5. Yes
6. No
7. I do not know
8. Prefer not to answer

**Core Section 9: Tobacco Use**

59. Have you smoked at least 100 cigarettes in your entire life? Do not include electronic cigarettes, herbal cigarettes, cigars, cigarillos, little cigars, pipes, bidis, kreteks, water pipes (hookahs) or marijuana, 5 packs = 100 cigarettes.

5. Yes
6. No
7. I do not know
8. Prefer not to answer

60. Do you now smoke cigarettes every day, some days, or not at all?

6. Every day
7. Some days
8. Not at all
9. I do not know
10. Prefer not to answer

61. During the past 12 months, have you stopped smoking for one day or longer because you were trying to quit smoking?

6. Yes
7. No
8. Not currently a smoker
9. I do not know
10. Prefer not to answer

62. How long has it been since you last smoked a cigarette, even one or two puffs?

6. Less than 1 month ago
7. More than 1 month but less than or equal to 3 months ago
8. More than 3 months but less than or equal to 6 months ago
9. More than 6 months but less than or equal to 1 year ago
10. More than 1 year but less than or equal to 5 years ago
11. More than 5 years but less than 10 years ago
12. 10 years or more
13. Never smoked regularly
14. I do not know
15. Prefer not to answer

63. Do you currently use chewing tobacco, snuff, or snus every day, some days, or not at all?

6. Every day
7. Some days
8. Not at all
9. I do not know
10. Prefer not to answer

**Core Section 10: Alcohol Consumption**

64. During the past 30 days, how many days per week or per month did you have at least one drink of any alcoholic beverage such as beer, wine, a malt beverage or liquor? (Select one)

6. _ _ Days per week
7. _ _ Days in past 30 days
8. No drinks in past 30 days
9. I do not know
10. Prefer not to answer

65. One drink is equivalent to a 12-ounce beer, a 5-ounce glass of wine, or a drink with one shot of liquor. During the past 30 days, on the days when you drank, about how many drinks did you drink on the average?

5. _ _ Number of drinks
6. No drinks in past 30 days
7. I do not know
8. Prefer not to answer

66. Considering all types of alcoholic beverages, how many times during the past 30 days did you have X [If A for #34 then, X= 5; if B for #34, X=4; if C or D for #34, X=4] or more drinks on an occasion?

5. _ _ Number of times
6. None
7. I do not know
8. Prefer not to answer

67. During the past 30 days, what is the largest number of drinks you had on any occasion?

4. _ _ Number of drinks
5. I do not know
6. Prefer not to answer

**Core Section 11: Exercise (Physical Activity)**

68.During the past month, other than your regular job, did you participate in any physical activities or exercises such as running/walking, weight lifting, playing sports, or other physical activities?

5. Yes
6. No
7. I do not know
8. Prefer not to answer

69. *[If A to #69]* Referring to the exercise list below, what type of physical activity or exercise did you spend the *most* time doing during the past month?

4. _ _ Specify from Physical Activity Coding List (below)

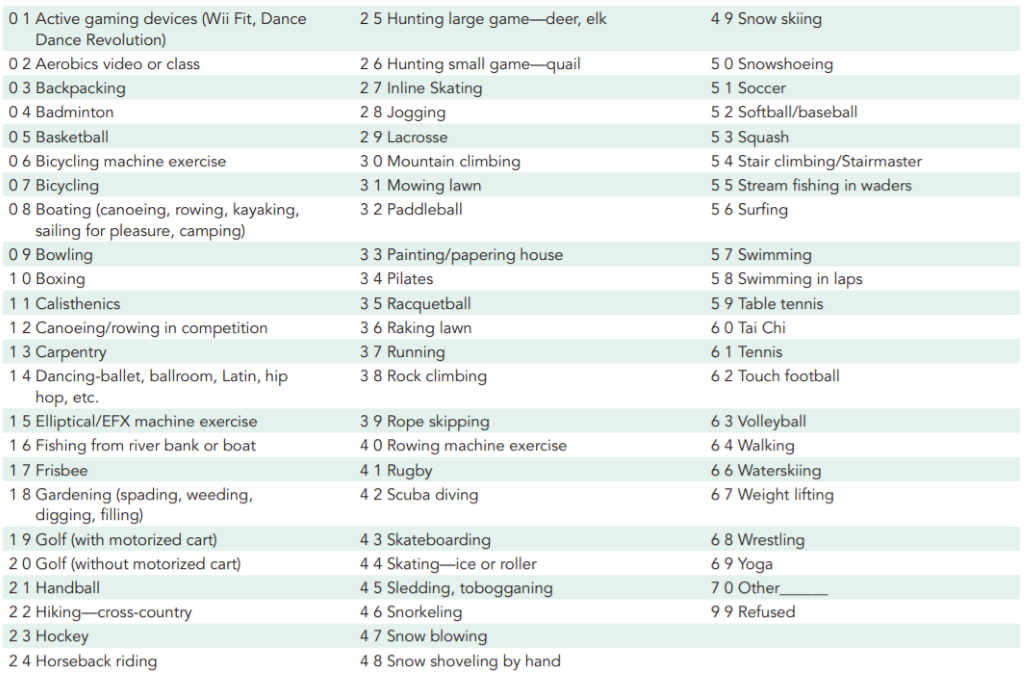

70. *[If A to #69]* How many times per week or per month did you take part in this activity during the past month?

5. _ _ Times per week
6. _ _ Times per month
7. I do not know
8. Prefer not to answer

71. *[If A to #69]* And when you took part in this activity, for how many minutes did you usually keep at it?

4. _ _ Minutes
5. I do not know
6. Prefer not to answer

72. *[If A to #69]* Referring to the exercise list, what other type of physical activity gave you the *next* most exercise during the past month?

5. _ _ _ Specify from physical activity list (below)
6. No other activity
7. I do not know
8. Prefer not to answer

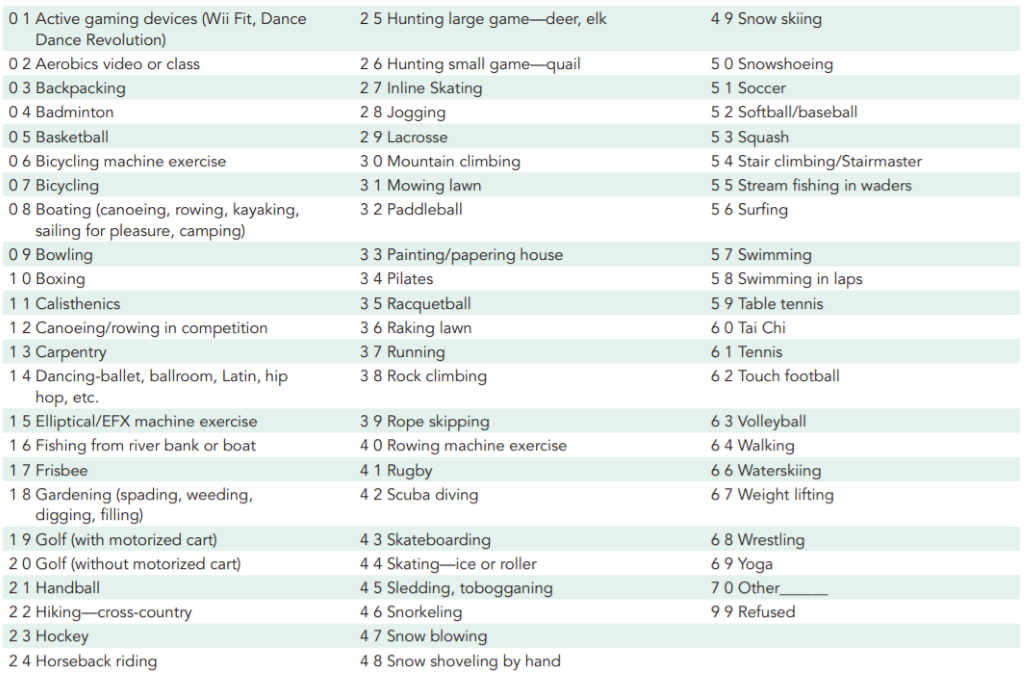

73. *[If A to #72]* How many times per week or per month did you take part in this activity during the past month?

5. _ _ Times per week
6. _ _ Times per month
7. I do not know
8. Prefer not to answer

74. *[If A to #72]* And when you took part in this activity, for how many minutes did you usually keep at it?

4. _ _ Minutes
5. I do not know
6. Prefer not to answer

75. *[If A to #69]* During the past month, how many times per week or per month did you do physical activities or exercises to strengthen your muscles?

6. _ _ Times per week
7. _ _ Times per month
8. Never
9. I do not know
10. Prefer not to answer

**Core Section 12: Fruits and Vegetables**

Now think about the foods you ate or drank during the past month, that is, the past 30 days, including meals and snacks.

76. Not including juices, how often did you eat fruit?

8. _ _ Day
9. _ _ Week
10. _ _ Month
11. Less than once a month
12. Never
13. I do not know
14. Prefer not to answer

77. Not including fruit-flavored drinks or fruit juices with added sugar, how often did you drink 100% fruit juice such as apple or orange juice? Do not include fruit-flavored drinks with added sugar like cranberry cocktail, Hi-C, lemonade, KoolAid, Gatorade, Tampico, and sunny delight. Include only 100% pure juices or 100% juice blends.

8. _ _ Day
9. _ _ Week
10. _ _ Month
11. Less than once a month
12. Never
13. I do not know
14. Prefer not to answer

78. How often did you eat a green leafy or lettuce salad, with or without other vegetables?

8. _ _ Day
9. _ _ Week
10. _ _ Month
11. Less than once a month
12. Never
13. I do not know
14. Prefer not to answer

79. How often did you eat any kind of fried potatoes, including French fries, home fries, or hash browns?

8. _ _ Day
9. _ _ Week
10. _ _ Month
11. Less than once a month
12. Never
13. I do not know
14. Prefer not to answer

80. How often did you eat any other kind of potatoes, or sweet potatoes, such as baked, boiled, mashed potatoes, or potato salad?

8. _ _ Day
9. _ _ Week
10. _ _ Month
11. Less than once a month
12. Never
13. I do not know
14. Prefer not to answer

81. Not including lettuce salads and potatoes, how often did you eat other vegetables?

8. _ _ Day
9. _ _ Week
10. _ _ Month
11. Less than once a month
12. Never
13. I do not know
14. Prefer not to answer

**Core Section 13: Vaccinations**

82. During the past 12 months, have you had either a flu vaccine that was sprayed in your nose or a flu shot injected into your arm?

5. Yes
6. No
7. I do not know
8. Prefer not to answer

83. *[If A to #83]* During what month and year did you receive your most recent flu vaccine that was sprayed in your nose or flu shot injected into your arm?

4. _ _ / _ _ _ _
5. I do not know
6. Prefer not to answer

84. Have you received a tetanus shot in the past 10 years?

7. Yes, received Tdap
8. Yes, received tetanus shot, but not Tdap
9. Yes, received tetanus shot but not sure what type
10. No, did not receive any tetanus shot in the past 10 years
11. I do not know
12. Prefer not to answer

85. Have you ever had a pneumonia shot also known as a pneumococcal vaccine?

5. Yes
6. No
7. I do not know
8. Prefer not to answer

86. Have you ever had the Human Papilloma virus vaccination or HPV vaccination?

5. Yes
6. No
7. I do not know
8. Prefer not to answer

87. *[If A to #87]* How many HPV shots did you receive?

5. _ _ Number of shots
6. All shots
7. I do not know
8. Prefer not to answer

88. Have you had a positive test for COVID-19 or coronavirus?

7. Yes
8. No
9. Test wasn’t able to provide results
10. Was not tested
11. I do not know
12. Prefer not to answer

**Please let us know how much you agree or disagree with the statements below.**

89. A vaccine developed for COVID-19 or coronavirus would be important for my health.

6. Strongly Disagree
7. Disagree
8. Neutral
9. Agree
10. Strongly Agree

90. A vaccine developed for COVID-19 or coronavirus would be effective.

6. Strongly Disagree
7. Disagree
8. Neutral
9. Agree
10. Strongly Agree

91. Being vaccinated for COVID-19 or coronavirus would be important for the health of others in my community.

6. Strongly Disagree
7. Disagree
8. Neutral
9. Agree
10. Strongly Agree

92. All vaccines offered by the government program in my community are beneficial.

6. Strongly Disagree
7. Disagree
8. Neutral
9. Agree
10. Strongly Agree

93. New vaccines carry more risks than older vaccines.

6. Strongly Disagree
7. Disagree
8. Neutral
9. Agree
10. Strongly Agree

94. The information I receive about vaccines from public health officials is reliable and trustworthy. Public health officials include the Illinois Department of Public Health, Centers for Disease Control and Prevention (CDC), and the United States Food and Drug Administration (FDA).

6. Strongly Disagree
7. Disagree
8. Neutral
9. Agree
10. Strongly Agree

95. The information I receive about vaccines from my doctor or healthcare provider is reliable and trustworthy.

6. Strongly Disagree
7. Disagree
8. Neutral
9. Agree
10. Strongly Agree

96. Getting vaccines is a good way to protect me from disease.

6. Strongly Disagree
7. Disagree
8. Neutral
9. Agree
10. Strongly Agree

97. Generally, I do what my doctor or health care provider recommends about vaccines.

6. Strongly Disagree
7. Disagree
8. Neutral
9. Agree
10. Strongly Agree

98. I would be concerned about serious adverse effects of a vaccine developed for COVID-19 or coronavirus.

6. Strongly Disagree
7. Disagree
8. Neutral
9. Agree
10. Strongly Agree

99. I would receive a vaccine developed for COVID-19 or coronavirus.

6. Strongly Disagree
7. Disagree
8. Neutral
9. Agree
10. Strongly Agree

**Module 1: Prediabetes**

100. Have you had a test for high blood sugar or diabetes within the past three years?

5. Yes
6. No
7. I do not know

101. Have you ever been told by a doctor or other health professional that you have pre-diabetes or borderline diabetes?

6. Yes
7. Yes, during pregnancy
8. No
9. I do not know

**Module 2: Diabetes**

102. *[If A to #32]* Are you now taking insulin?

5. Yes
6. No
7. I do not know

103 *[If A to #32]* About how often do you check your blood for glucose or sugar?

8. _ _ Times per day
9. _ _ Times per week
10. _ _ Times per month
11. _ _ Times per year
12. Never
13. I do not know

104 *[If A to #32]* Including times when checked by a family member or friend, about how often do you check your feet for any sores or irritations?

9. _ _ Times per day
10. _ _ Times per week
11. _ _ Times per month
12. _ _ Times per year
13. No feet
14. Never
15. I do not know

105 *[If A to #32]* About how many times in the past 12 months have you seen a doctor, nurse, or other health professional for your diabetes?

5. _ _ Number of times
6. None
7. I do not know

106 *[If A to #32]* About how many times in the past 12 months has a doctor, nurse, or other health professional checked your A1C?

6. _ _ Number of times
7. None
8. Never heard of A1C test
9. I do not know

107 *[If A to #32]* About how many times in the past 12 months has a health professional checked your feet for any sores or irritations?

5. _ _ Number of times
6. None
7. I do not know

108 *[If A to #32]* When was the last time you had an eye exam in which the pupils were dilated, making you temporarily sensitive to bright light?

1. Within the past month
2. More than 1 month but less than or equal to 12 months ago
3. More than 1 year but less than 2 years ago
4. 2 or more years ago
5. I do not know
6. Never

109. *[If A to #32]* Has a doctor ever told you that diabetes has affected your eyes or that you had retinopathy?

5. Yes
6. No
7. I do not know

110. *[If A to #32]* Have you ever taken a course or class in how to manage your diabetes yourself?

5. Yes
6. No
7. I do not know

**Module 9: Breast and Cervical Cancer Screening**

111. [*If B to #34*] Have you ever had a mammogram?

5. Yes
6. No
7. I do not know

112. [*If A to #112*] How long has it been since you had your last mammogram?

1. Within the past year
2. More than 1 year but less than or equal to 2 years ago
3. More than 2 years but less than or equal to 3 years ago
4. More than 3 years but less than 5 years ago
5. 5 or more years ago
6. I do not know

113. *[If B to #34]* Have you ever had a Pap test?

5. Yes
6. No
7. I do not know

114.*[If A to #114]* How long has it been since you had your last Pap test?

1. Within the past year
2. More than 1 year but less than or equal to 2 years ago
3. More than 2 years but less than or equal to 3 years ago
4. More than 3 years but less than 5 years ago
5. 5 or more years ago
6. I do not know

115. *[If A to #115]* An H.P.V. test is sometimes given with the Pap test for cervical cancer screening. Have you ever had an H.P.V. test?

5. Yes
6. No
7. I do not know

116. [*If A to #116*] How long has it been since you had your last H.P.V. test?

1. Within the past year
2. More than 1 year but less than or equal to 2 years ago
3. More than 2 years but less than or equal to 3 years ago
4. More than 3 years but less than 5 years ago
5. 5 or more years ago
6. I do not know

117. [*If B to #34*] Have you had a hysterectomy?

5. Yes
6. No
7. I do not know

**Module 12: Colorectal Cancer Screening**

118. A blood stool test is a test that may use a special kit at home to determine whether the stool contains blood. Have you ever had this test using a home kit?

5. Yes
6. No
7. I do not know
8. Prefer not to answer

119. *[If A to #119]* How long has it been since you had your last blood stool test using a home kit?

1. Within the past year (anytime less than 12 months ago)
2. Within the past 2 years (1 year but less than 2 years ago)
3. Within the past 3 years (2 years but less than 3 years ago)
4. Within the past 5 years (3 years but less than 5 years ago)
5. 5 or more years ago
6. I do not know
7. Prefer not to answer

120. Sigmoidoscopy and colonoscopy are exams in which a tube is inserted into the rectum to view the colon for signs of cancer or other health problems. Have you ever had either of these exams?

5. Yes
6. No
7. I do not know
8. Prefer not to answer

121. *[If A to #121]* For a sigmoidoscopy, a flexible tube is inserted into the rectum to look for problems. A colonoscopy is similar, but uses a longer tube, and you are usually given medication through a needle in your arm to make you sleepy and told to have someone else drive you home after the test. Was your most recent exam a sigmoidoscopy or a colonoscopy?

5. Sigmoidoscopy
6. Colonoscopy
7. I do not know
8. Prefer not to answer

122. *[If A to #121]* How long has it been since you had your last sigmoidoscopy or colonoscopy?

9. Within the past year (anytime less than 12 months ago)
10. Within the past 2 years (1 year but less than 2 years ago)
11. Within the past 3 years (2 years but less than 3 years ago)
12. Within the past 5 years (3 years but less than 5 years ago)
13. Within the past 10 years (5 years but less than 10 years ago)
14. 10 or more years ago
15. I do not know
16. Prefer not to answer

**Module 14: Healthcare Access**

123. [*If A to #13*] What is the primary source of your health care coverage?

11. A plan purchased through an employer or union (including plans purchased through another person’s employer)
12. A plan that you or another family member buys
13. Medicare
14. Medicaid or other state program
15. TRICARE (formerly CAMPUS), VA, or Military
16. Alaska Native, Indian Health Service, Tribal Health Services
17. Some other source
18. None (no coverage)
19. I do not know
20. Prefer not to answer

**Module 22: Adverse Childhood Experiences** The following section will ask sensitive questions about depression, substance abuse, domestic violence, and sexual abuse within your family unit. We acknowledge that these questions may be difficult to answer and understand if certain questions will be answered with “Prefer not to answer”. We would also like to let you know that if you would like resources for mental health, substance abuse, domestic violence, and sexual abuse, please contact the UIC Counseling Center at (312) 996-3490 or visit <https://counseling.uic.edu/>

124. Now, looking back before you were 18 years of age, did you live with anyone who was depressed, mentally ill, or suicidal?

5. Yes
6. No
7. I do not know
8. Prefer not to answer

125. Did you live with anyone who was a problem drinker or alcoholic?

5. Yes
6. No
7. I do not know
8. Prefer not to answer

126. Did you live with anyone who used illegal street drugs or who abused prescription medications?

5. Yes
6. No
7. I do not know
8. Prefer not to answer

127. Did you live with anyone who served time or was sentenced to serve time in a prison, jail, or other correctional facility?

5. Yes
6. No
7. I do not know
8. Prefer not to answer

128. Were your parents separated or divorced?

6. Yes
7. No, parents still married
8. Parents never married
9. I do not know
10. Prefer not to answer

129. How often did your parents or adults in your home ever slap, hit, kick, punch, or beat each other up?

6. Never
7. Once
8. More than once
9. I do not know
10. Prefer not to answer

130. Not including spanking (before age 18), how often did a parent or adult in your home ever hit, beat, kick, or physically hurt you in any way?

6. Never
7. Once
8. More than once
9. I do not know
10. Prefer not to answer

131. How often did a parent or adult in your home ever swear at you, insult you, or put you down?

6. Never
7. Once
8. More than once
9. I do not know
10. Prefer not to answer

132. How often did anyone ever touch you sexually without consent?

6. Never
7. Once
8. More than once
9. I do not know
10. Prefer not to answer

133. How often did anyone try to make you touch them sexually?

6. Never
7. Once
8. More than once
9. I do not know
10. Prefer not to answer

134.How often did anyone force you to have sex?

6. Never
7. Once
8. More than once
9. I do not know
10. Prefer not to answer

**Module 25: Marijuana Use**

135. During the past 30 days, on how many occasions did you use marijuana or cannabis?

5. _ _ Number of days
6. None
7. I do not know
8. Prefer not to answer

136. *[If A to #136]* During the past 30 days, which one of the following ways did you use marijuana the most often?

9. Smoke it (for example, in a joint, bong, pipe, or blunt)
10. Eat it (for example, in brownies, cakes, cookies, or candy)
11. Drink it (for example, in tea, cola, or alcohol)
12. Vaporize it (for example, in an e-cigarette-like vaporizer or another vaporizing device)
13. Dab it (for example, using waxes or concentrates)
14. Use it some other way
15. I do not know
16. Prefer not to answer

137. *[If A to #136]* When you used marijuana or cannabis during the past 30 days, was it usually

6. For medical reasons (to treat or decrease symptoms of a health condition)
7. For non-medical reasons (like to have fun or fit in)
8. For both medical and non-medical reasons
9. I do not know
10. Prefer not to answer

**Module 29: Sexual Orientation and Gender Identity**

138. Which of the following best represents how you think of yourself?

7. Gay
8. Straight, that is not gay
9. Bisexual
10. Something else
11. I do not know
12. Prefer not to answer

139 Do you consider yourself to be transgender?

7. Yes, transgender, male to female
8. Yes, transgender, female to male
9. Yes, transgender, gender nonconforming
10. No
11. I do not know
12. Prefer not to answer

**That is the end of our questionnaire! You will receive a link to an Amazon.com electronic gift card for $5 to your primary email address.**

**Please feel free to contact the study team at 312-413-2873 if you have any questions.**

**Thank you for your participation**

**Appendix 3**

*Includes a breakdown of the demographics of eligible health professional students by health science college*

| **UIC Health Science College** | **Demographics** | | |
| --- | --- | --- | --- |
|  | **Enrolled, number^1^** | **Gender (%)^2^** | **Underrepresented minorities (%)^3^** |
| **College of Applied Health Sciences**  BS: Health Information Management  BS: Health Information Management (Online)  BS: Nutrition-Coordinated Program  MS: Nutrition  DPT: Physical Therapy  MS: Occupational Therapy | 462 | Male: 21.6%  Female: 78.4% | 30.1% |
| **College of Medicine**  MD: Medicine - Chicago  MD: Medicine - Peoria  MD: Medicine - Rockford  MD: Medicine -Urbana | 1,264 | Male: 51.2%  Female: 48.8% | 19.4% |
| **College of Dentistry**  DMD: Dental Medicine  DMD-AS: Dental Medicine-Advanced Standing | 378 | Male: 44.2%  Female: 55.8% | 20.1% |
| **College of Nursing**  BSN: Registered Nurse to Bachelor of Nursing - Chicago  BSN: Nursing - Chicago  BSN: Nursing - Springfield  BSN: Nursing – Urbana  MS: Nursing - Chicago  MS: Nursing - Urbana  DNP: Nursing Practice -Chicago  DNP: Nursing Practice - Peoria  DNP: Nursing Practice – Quad Cities  DNP: Nursing Practice - Springfield  DNP: Nursing Practice - Urbana  DNP: Nursing Practice - Rockford | 1,398 | Male: 12.8%  Female: 87.2% | 28.4% |
| **College of Pharmacy**  PHARMD: Pharmacy - Chicago  PHARMD: Pharmacy - Rockford | 742 | Male: 40.2%  Female: 59.8% | 16.3% |
| **School of Public Health**  MHA: Healthcare Administration  MPH: Community Health Sciences (Online)  MPH: Community Health Sciences  MPH: Public Health Informatics  MPH: Health Policy & Administration (Online)  MPH: Health Policy & Administration  MPH: Environmental & Occupational Health Sciences  MPH: Biostatistics  MPH: Epidemiology | 396 | Male: 23.0%  Female: 77.0% | 31.3% |
| **Jane Addams College of Social Work**  MSW: Social Work | 478 | Male: 15.9%  Female: 84.1% | 44.6% |
| **TOTAL:** | 5,118 | Male: 30.4%  Female: 69.6% | 26.7% |

Data Source: Fall 2020 UIC OIR Census file

1.“Number” includes the total students in health professional programs at each health science college.

2.“Gender” includes the percentage of male and female students at each health science college across health professional programs (census did not provide another option for gender).

3.“Underrepresented minorities” measures the total percentage of Underrepresented Minority (URM) students at each health science college across health professional programs. Underrepresented minority students include the following groups: American Indian Alaskan Native (AIAN), Black/African American, Hispanic, and Multi-Race.
